# Infant- and child-level predictors of mortality in low-resource settings: the WHO Child Mortality Risk Stratification Multi-Country Pooled Cohort

**DOI:** 10.1101/2024.07.06.24309988

**Authors:** The WHO Risk Stratification Working Group (WHO-RSWG), Catherine Schwinger

## Abstract

**Background:** Despite impressive reductions in overall global child mortality, rates of decline have slowed during the last decade. Current guidelines for the care of at-risk children in low-resource settings mostly focus on broad clinical syndromes or undernutrition rather than children’s individual contextualized risk. We aimed to identify readily assessable child-level characteristics that can predict child mortality risk in a range of community and healthcare settings in high-burden settings.

**Methods:** We analysed pooled data from 33 cohorts including 75,287 children under five years of age living in low resource settings to estimate the absolute risks of death associated with risk exposures separately and combined with anthropometry. Children were grouped according to population types studied: general population (GP), selected on anthropometric criteria (A-S), and selected on the presence of illness (I-S).

**Findings:** During a total of 69,085 child-years of follow-up in the pooled dataset, 2,805 (3.7%) children died. Age <24 months, low anthropometry, preterm birth, low birthweight, and absence of breastfeeding were each associated with increased mortality: risks were additive and declined with increasing age. However, overall mortality and the association between child-level characteristics and mortality differed according to the type of study population and child age.

**Interpretation:** Risk assessments combining individual child-level characteristics including anthropometry can enable programmes to identify children at high and lower risk of mortality and, thereafter, differentiate care accordingly. Such a strategy may reduce mortality and optimise health system efficiency and effectiveness.

## INTRODUCTION

Global rates of under-5 child mortality (U5M) have declined from 93 to 38 deaths per 1,000 live births between 1990 and 2021.^1^ However, since 2010, U5M reductions have slowed; 53 countries will not meet the 2030 UN Sustainable Development Goals (SDG) U5M target.^2^ More than 80% of global U5M occurs in sub-Saharan Africa and South Asia, with significant sub-national heterogeneity.^3,4^ Mortality also varies markedly by age, with a third of U5M in some settings occurring within the first three days of life^5^ and almost half occurring in the first month.^2^

Low birth weight (LBW), including from preterm birth (PTB), is associated with half of neonatal deaths and confers life-long health risks.^2^ Additionally, undernutrition in all its forms contributes to 45% of U5M.^6^ Chronic disease and disability, maternal illiteracy, poor housing conditions, and limited access to healthcare are less often assessed, but may all directly or indirectly be associated with child mortality.^7^ Focusing on interventions that target underlying pathways associated with these risks may offer opportunity to reduce mortality. Importantly, however, while a focus on higher-risk children may avert some mortality, identifying lower risk children may provide opportunities for de-escalation of care and to optimize available resources while improving care for those at highest risk. While prognostic scores have been devised among hospitalised children to identify those at high risk, they are rarely used in practice in LMICs.^8,9^

We estimated absolute and relative mortality risks associated with separate and combined assessable, child-level characteristics among children aged 0-59 months through an individual data pooled analysis of the WHO Child Mortality Risk Stratification Multi-Country Pooled Cohort (WHO CMRS).

## METHODS

### Study design

We pooled data from individual children enrolled in observational or randomized controlled trials in LMICs (Table S1). The criteria for inclusion of a dataset were 1) documentation of age, weight, vital status, date of death, and 2) at least two observations per participant under the age of 60 months. Details of the study selection procedure are reported fully in the cohort profile.^10^

### Data management

Thirty-three datasets were included and pooled into a single database with consistent variable names and structures. Data cleaning included checking for missing data, duplicates, and outliers^10^. Anthropometric measurements were defined as outliers if mid-upper arm circumference (MUAC) >300mm or <55mm, weight-for-age (WAZ) >10 or <-10, length <35cm, length- or height-for-age (HAZ) >10 or <-12, weight-for-length or -height (WHZ) >20 or <-10 and values set to missing. As we used anthropometric measurements as categorical variables in analyses, and an extreme value would likely be in the severe deficit category (Z-score <-3), we did not apply more strict cleaning criteria.

### Indicator assessment and definition

In all analyses, the outcome was defined as death within the respective observation period. If the vital status of a child was recorded but age at death was missing, we estimated the age at death by adding half of the median observation period of the study to the age at the last study contact point for that child.

Child-level characteristics were chosen as risk exposure variables for analysis based on availability across datasets. Weight and height measurements were converted to Z-scores using WHO Child Growth Standards.^11^ We defined a moderate anthropometric deficit as Z-score in WAZ, HAZ, or WHZ <-2 and ≥-3, and a severe deficit as Z-score <-3. MUAC was included as an absolute measure (mm). The WHO guidelines recommend the cut-off for moderate and severe deficits for MUAC at 125mm and 115mm for children aged 6-59 months.^12^ For infants <6 months of age, we also applied cut-offs of 120mm and 110mm for moderate and severe deficits.^12^

All other child-level characteristics were dichotomized. Breastfeeding was categorized as yes if the mother reported any breastfeeding in the 30-day period before assessment.^10^ LBW was defined as <2500g, PTB as gestational age <37 weeks. Due to limitations in gestational assessments, we did not include a variable for small for gestational age (SGA). We captured illness by including any reported episode of diarrhoea within the prior 14 days, and study physician diagnoses of pneumonia or caretaker reports of WHO symptom classifications in the last 14 days for lower respiratory tract infections (LRTI). Severe illness was defined as being admitted to hospital or having any WHO Integrated Management of Childhood Illness (IMCI) danger sign, i.e., not able to drink or breastfeed, convulsions, lethargic or unconscious^13^. The IMCI danger sign of vomiting everything was not consistently available for inclusion in the pooled dataset.

Each study was classified into one of three population types based on the assumption that associations between risk exposures and mortality would differ between subsets of children: (1) “General population” (GP) if enrolment to the original study was not based on an anthropometric deficit or the presence of an illness, (2) Anthropometry-Selected (A-S) if children were enrolled based on an anthropometric deficit, or (3) Illness-Selected (I-S) if children were enrolled on the basis of signs or symptoms of an illness (potentially in addition to other eligibility criteria)^10^.

### Statistical analysis

All statistical analyses were done using R version 4.3.1 (R Foundation for Statistical Computing, Vienna, Austria, 2023). To estimate the probability of dying within an observation period, we used multiple generalized linear mixed effects regression (glmer) models of the binomial distribution family and logit link function. The dependent variable of these models was whether a child died during the observation period, and all models included child and study as random intercepts. As the time intervals between observations affect the possibility of capturing death, we also included the log transformed time in months between each visit as an offset.

We built separate models for each exposure; all models included age at time of visit. From these models, we calculated odds ratios (see supplementary material) and the predicted absolute risk of death per child-month. We report these according to age groups 0-5, 6-11, 12-23, and 24-59 months. In addition, we graphically display the predicted absolute mortality risk over the age range of 0-24 months. All models included an interaction term between age at the time of visit and the other independent variable. To account for differences in the completeness of data, missing observations from independent variables were included as a distinct category, such that the risk of dying was separately calculated for children with missing values.

To estimate the association of not being breastfed, LBW, or PTB, combined with an anthropometric deficit, we estimated the absolute risk using the same statistical approach noted above. We estimate the risk for none, only one of the exposures (e.g., underweight but breastfed) and both together (e.g., underweight AND not breastfed). Only children with information on both exposures were used in the analyses. Data availability for each study is detailed in the cohort profile.^10^

All analyses were run separately for the three population categories: GP, A-S, or I-S. In additional analyses, we report absolute risks of death only for children with death age available (Table S2). We decided a priori to report only those estimates where there were more than 20 observations within each age-specific risk exposure group.

### Ethics

The funders of the studies had no role in study design, data collection, data analysis, data interpretation, or writing of the report. All principal investigators contributing data signed an agreement with WHO for pooling and use of anonymous study data for pre-defined analyses. All study investigators confirmed that each included study had appropriate ethical review approval and that informed consent was obtained from all participants. The WHO Ethics Review Committee granted an exemption in March 2022 (ERC.0003745). The pooled data are kept on a secure server at the University of Bergen, Norway with restricted access.

## RESULTS

### Pooled dataset

Of 35 datasets received, two did not meet the inclusion criteria (Figure 1), and data from 33 studies were included and pooled. In total, 4·5% observations were excluded from analyses (i.e., 949 duplicate observations, 22,342 observations without age at a single observation timepoint, 160 observations occurring at age >60 months, and 1,047 observations without information on sex).^10^ The sample sizes of included studies ranged from 247 to 7,956 children. The final pooled dataset included individual-level data from 75,287 children aged 0-5 years and 546,459 observations. There were 23 studies classified as GP (49,740 children), six studies classified as A-S (14,759 children), and four studies classified as I-S (10,788 children) (Figure 1).

**Figure 1:**
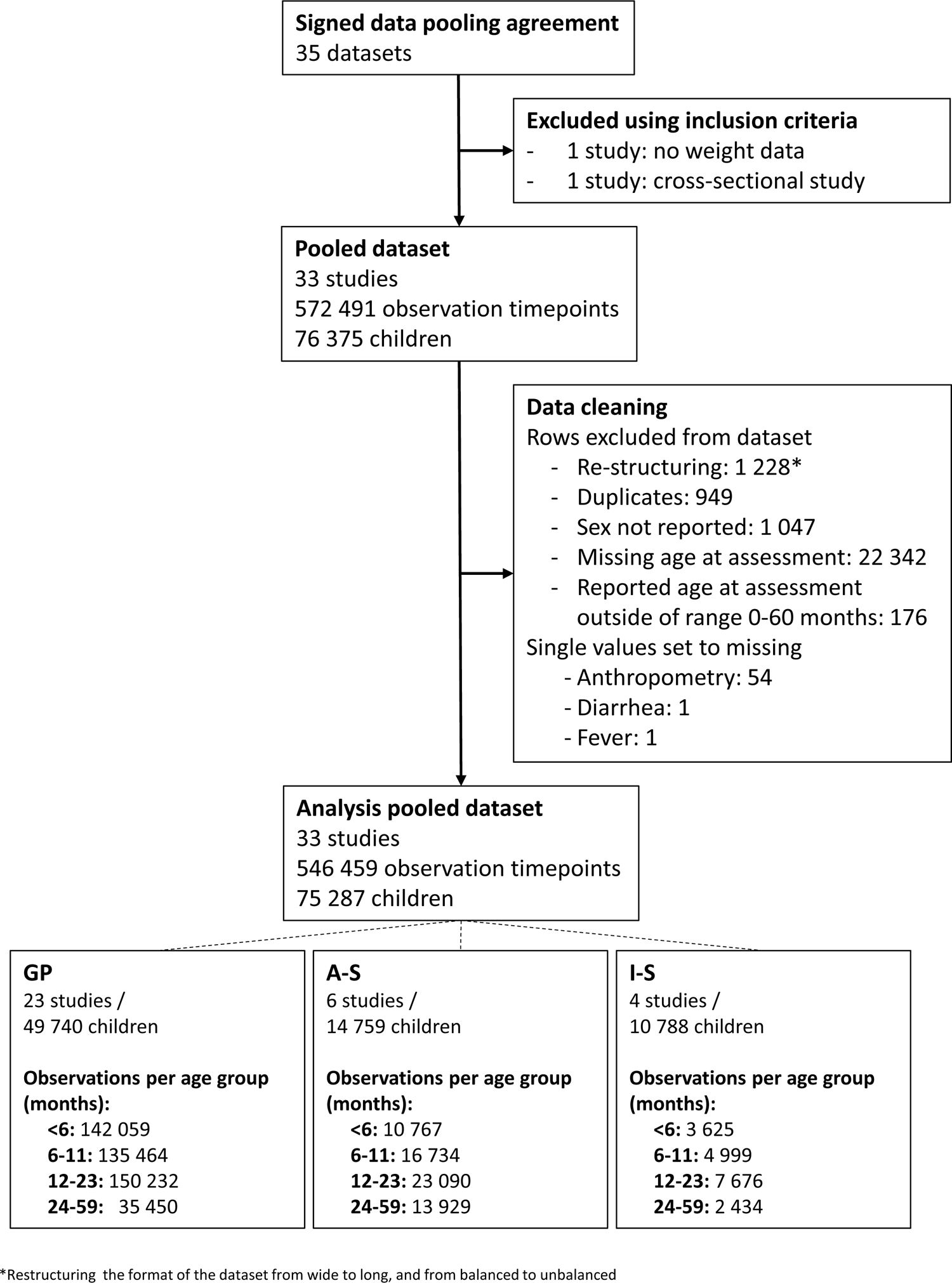
Study flow

Baseline characteristics of children differed between the three population types (Table 1). Children in GP studies were younger at enrolment (median age of one month as compared to 13·6 months (A-S) and 10·7 months (I-S)). In studies in which birthweight and gestational age were collected, the proportion of children without information on birthweight was 6% among GP studies (1,659/28,326 children), 14% among A-S studies (174/1,286), and 10% among I-S studies (956/9,515). The proportion of missing data on PTB was 2% in GP studies (574/28,274 children) and 0·7% in I-S studies (47/6,414 children). None of the A-S studies had information on PTB. The proportion of missing data increased with age; among children aged 12-23 months, this information was not available for 24-39% of the children (Supplementary Table S3). The anthropometric values among children in A-S studies at time of first measurement were lower than in GP and I-S studies. Symptoms and signs of illness were more common among children in I-S studies. Median maternal HIV prevalence also differed significantly by study type (Table 1).

**Table 1:** Characteristics of children and their mothers in the pooled dataset according to enrolment criteria groups (general populations (GP), anthropometry-selected (A-S) and illness-selected (I-S) populations)

| Characteristic<br>(where data available) | GP<sup>1</sup> |  | A-S<sup>1</sup> |  | I-S<sup>1</sup> |  |
| --- | --- | --- | --- | --- | --- | --- |
|  | N<sup>2</sup> | % /<br>median (IQR) | N<sup>2</sup> | % /<br>median (IQR) | N<sup>2</sup> | % /<br>median (IQR) |
| **Maternal characteristics at enrolment** |  |  |  |  |  |  |
| Mother alive | 19,702 | 99.8% | 12,467 | 98.2% | 10,721 | 99.0% |
| Mother primary caretaker | 15,052 | 98.1% | 12,866 | 94.6% | 9,444 | 93.9% |
| Maternal age, years | 42,549 | 24 (20, 29) | 2,079 | 23 (21, 25) | 10,78 | 26 (22, 30) |
| Maternal education | 34,622 |  | 5,941 |  | 7,472 |  |
| no education |  | 35% |  | 37% |  | 17% |
| primary education |  | 25% |  | 10% |  | 25% |
| secondary or higher education |  | 40% |  | 53% |  | 58% |
| Mother HIV positive | 10,872 | 0.2% | 4,418 | 14.6% | 1,517 | 18.5% |
| **Child's characteristics at enrolment** | **N<sup>2</sup>** |  | **N<sup>2</sup>** |  | **N<sup>2</sup>** |  |
| Child's sex, male | 49,740 | 50.5% | 14,759 | 42.8% | 10,788 | 55.6% |
| Child age, months | 49,740 | 1.0 (0, 7.8) | 14,759 | 13.6 (8.3, 22.6) | 10,788 | 10.7 (4.7, 18.7) |
| Low birthweight, <2500g | 26,667 | 13.3% | 1,112 | 5.0% | 8,559 | 7.8% |
| Birthweight, g | 26,667 | 3000 (2700, 3400) | 1,112 | 3000 (2900, 3400) | 2,145 | 3000 (2500, 3500) |
| Preterm birth, <37 weeks of gestation | 27,700 | 15.7% | N/A | N/A | 6,367 | 4.3% |
| WAZ | 42,636 | -0.9 (-1.7, -0.1) | 14,754 | -2.9 (-3.6, -2.3) | 10,767 | -1.4 (-2.8, -0.4) |
| HAZ | 41,251 | -1.1 (-1.9, -0.3) | 14,747 | -2.5 (-3.5, -1.7) | 10,721 | -1.2 (-2.4, 0.1) |
| WHZ | 39,853 | -0.3 (-1.2, 0.7) | 14,168 | -2.2 (-2.8, -1.6) | 10,665 | -1.3 (-2.6, -0.1) |
| MUAC, mm | 33,423 | 120 (104, 135) | 14,717 | 118 (110, 123) | 10,762 | 128 (114, 141) |
| **Reported illness or feeding practice at any visit** | **Obs<sup>2</sup>** |  | **Obs<sup>2</sup>** |  | **Obs<sup>2</sup>** |  |
| Acute diarrhoea<sup>3</sup> | 313,381 | 7·6% | 53,279 | 30·2% | 18,672 | 43·7% |
| Acute lower respiratory tract infection<sup>4</sup> | 150,874 | 3·2% | N/A | N/A | 18,671 | 40·1% |
| Severe illness<sup>5</sup> | 242,980 | 1·2% | 36,476 | 9·6% | 17,461 | 28·4% |
| Any breastfeeding<sup>6</sup> |  |  |  |  |  |  |
| at age 0-5 months | 92,024 | 98·9% | 7,947 | 98·8% | 3,622 | 91·2% |
| at age 6-23 months | 202,644 | 87·0% | 25,314 | 81·6% | 12,673 | 70·6% |
| at age 24-59 months | 21,306 | 18·8% | 7,521 | 16·0% | 2,423 | 17·1% |

<sup>1</sup> We categorized studies on the basis of their enrolment criteria, where GP = study cohorts where enrolment was not based on anthropometry or the presence of illness, A-S = study cohorts where children were enrolled on the basis of an anthropometric deficit, I-S = study cohorts where children were enrolled on the basis of the presence of illness using study specific definitions.
<sup>2</sup> N = total number of children with data available for this variable, Obs = total number of observations available for this variable
<sup>3</sup> Any diarrheal episode reported by a clinician or caregiver within the last 24 hours, 3, 7 or 14 days.
<sup>4</sup> Pneumonia or acute lower respiratory tract infection reported by a clinician or caregiver within the last 14 days.
<sup>5</sup> Any of the following: being admitted to hospital, not being able to drink or breastfeed, having had convulsions, being lethargic or unconscious.
<sup>6</sup> Any breastfeeding within the past 14 days reported by caregiver at the study visit.
Abbreviations: HAZ = height-for-age Z-score (includes length-for-age Z-scores); IQR = interquartile range; MUAC = mid-upper arm circumference; N = number of children; Obs = number of observations, WAZ = weight-for-age Z-score; WHZ = weight-for-height Z-score (includes weight-for-length Z-scores).

Follow-up duration and frequency varied between studies: median (IQR) follow-up time per child was 17 (11,23), 3 (1,6), and 6 (5·9,6·2) months, in the GP, A-S and I-S populations, respectively. The median (IQR) interval between visits was 1 (0·9,1·3) month in the GP population, 0·5 (0·4,0·9) months in the A-S population, and 3 (1·6,6) months in the I-S population. Fourteen of the 23 GP studies were birth cohorts (enrolling up to 17 days after birth) including 25,992 of 49,739 children (52%); only one of six A-S studies (2,079/14,760 children; 14%) and none of four I-S studies enrolled from birth. See Table S1 and the cohort description.^10^

There were 2,805 deaths during a total of 69,085 child-years of follow up. Death occurred in 2·5% of children in the GP population (1,238/49,740), 2·5% in the A-S population (366/14,759) and 11% in the I-S population (1,201/10,788). Age at time of death was documented for 2,660 (95%) children.

### Mortality risks in the GP population

Age was a strong predictor of death, with the highest absolute risk of death within the first months after birth, declining with increasing age (see Figure 2). The risk of death increased with lower anthropometric status at all ages and for all anthropometric indices (Figure 2). The mortality risks in the age groups <6, 6-11, 12-23, and 24-59 months can be found in Supplementary Tables S3 and S4. The highest absolute mortality risks were among those with very low WAZ and youngest age.

**Figure 2:**
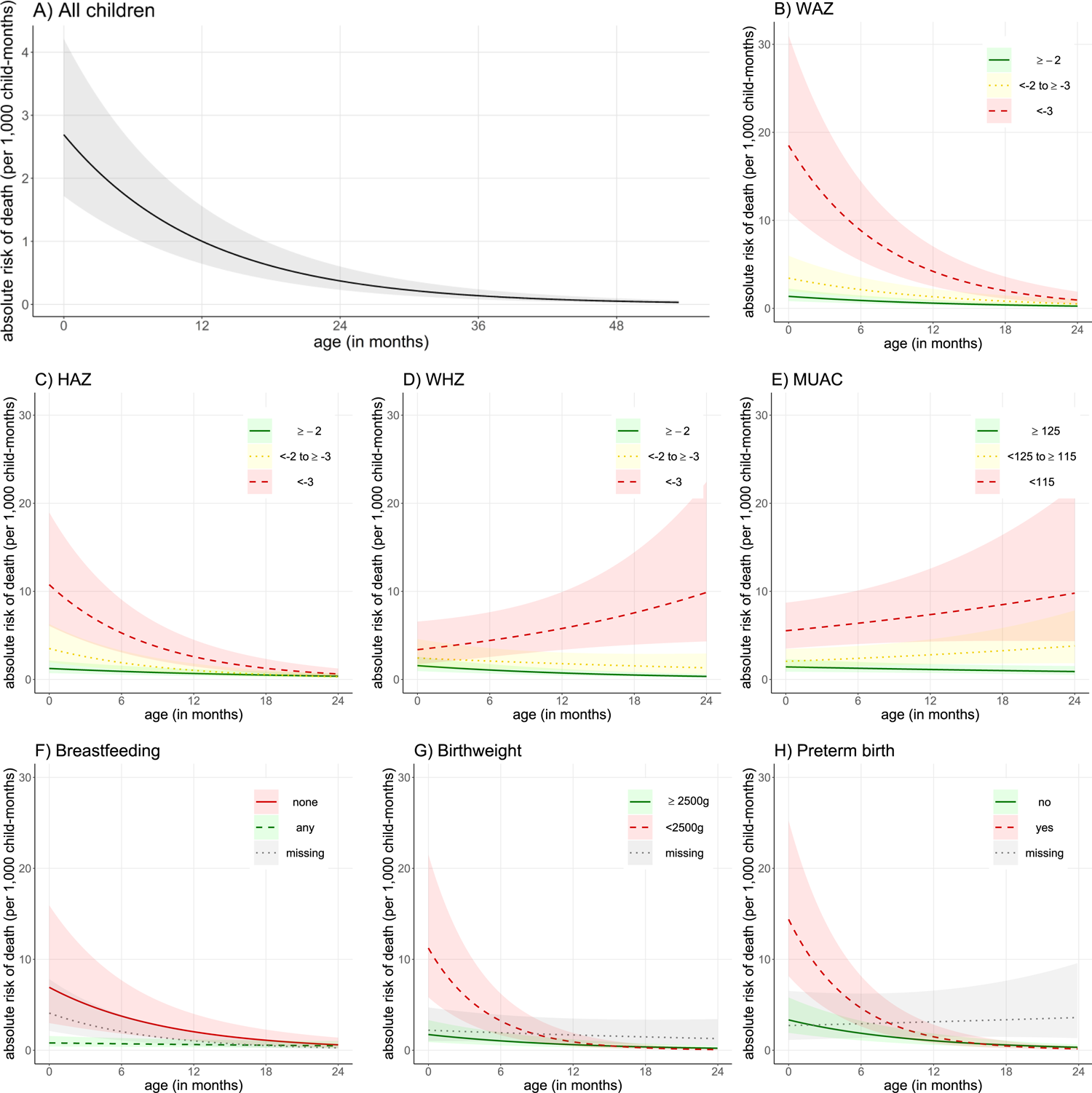
The absolute predicted risk of death in GP children 0-24 months of age according to A) age; B) weight-for-age Z-score (WAZ), C) length/height-for-age Z-score (HAZ), D) weight-for-length/height Z-score (WHZ), E) mid-upper arm circumference in mm (MUAC), F) breastfeeding practices, G) birthweight, and H) age of gestation at birth Reported symptoms of diarrhea or LRTI did not further discriminate mortality risks (Supplementary Table S3). Very few children were reported to have any severe illness as defined. Infants who were reported as not breastfeeding had a higher absolute mortality risk as compared to those who were receiving any breastfeeding (Figure 2). The mortality risk associated with not breastfeeding was highest in younger children and decreased gradually with increasing age. In the age group 24-59 months, there were few children still being breastfed with no differences in absolute mortality risk by breastfeeding status (Supplementary Table S3). The absolute mortality risk was increased among LBW or PTB infants and remained elevated until about one year of age (Figure 2).

However, the age gradient was not apparent for MUAC, where the absolute mortality risk was relatively stable between birth and 24 months (Figure 2). Applying modified cut-offs for moderate and severe deficits among children aged 0-5 months (i.e., 120 and 110mm) did not substantially change the risk estimates (Supplementary Table S3). The mortality risk associated with a deficit in WAZ by stunting status changed with age (Supplementary Table S5). Among children with low WAZ aged less than 12 months, concurrent stunting increased the mortality risk; among those with low WAZ and aged 12 months or more, concurrent stunting was associated with lower mortality risk.

In analyses that combined risk exposures, the absolute risk of mortality in children with even a moderate anthropometric deficit (<-2 Z-scores) was further increased when there was a history of LBW, PTB, or no reported breastfeeding (Table 2 and Figure 3).

**Figure 3.**
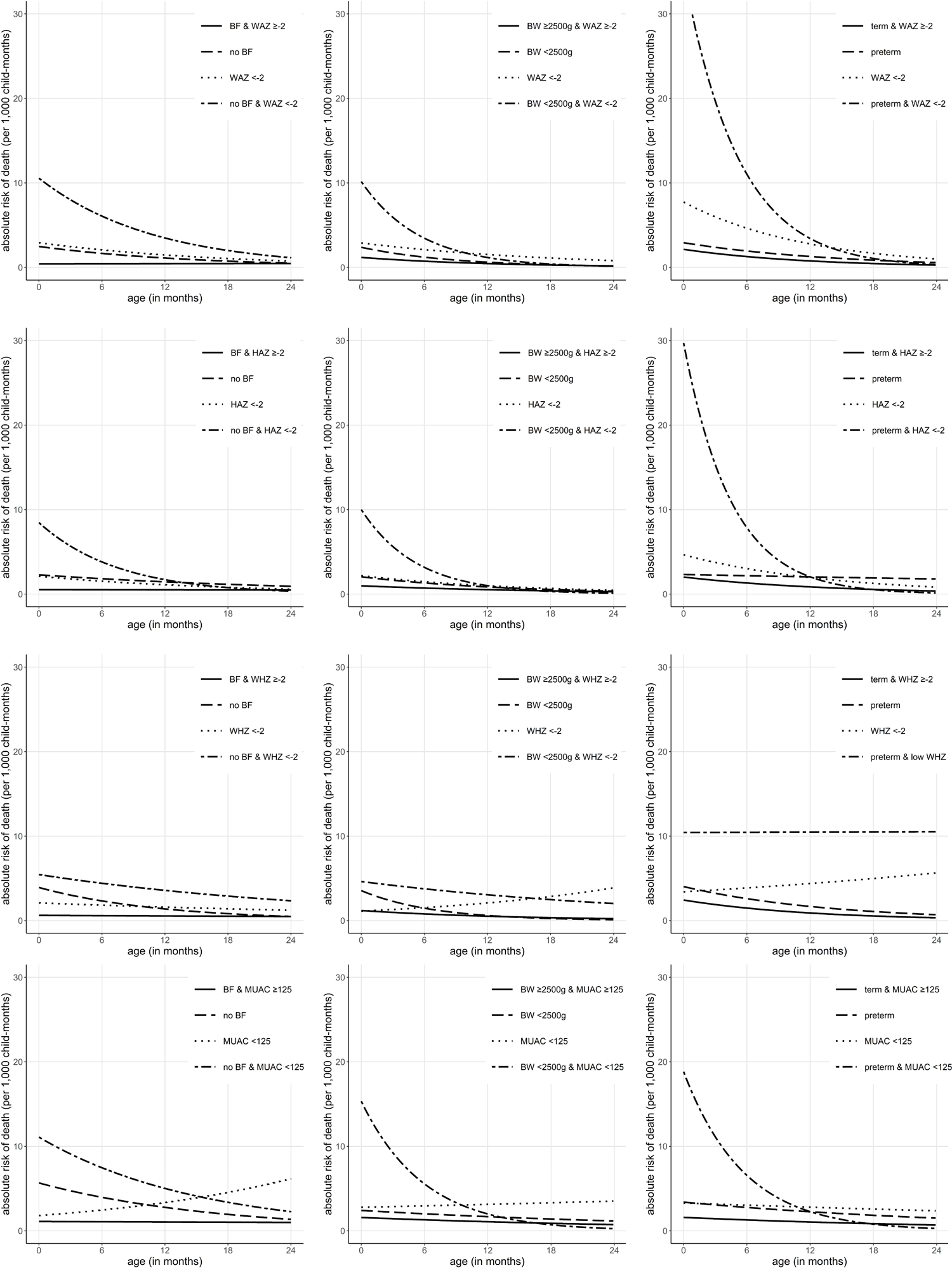
The absolute risk of death in GP children 0-24 months of age with combined risk exposures: either not breastfeeding, low birthweight, or being born preterm, combined with low weight-for-age (WAZ), low height-for-age (HAZ), low weight-for-height (WHZ) or low mid-upper arm circumference (MUAC). Analyses are restricted to children where data on both indicators are available (see also Table 2)

**Table 2:** Absolute risk of death per 1,000 child months by age group and by combinations of exposures in General Population (GP) cohorts, i.e., enrolment not on the basis of anthropometric deficit or the presence of illness.

|  | <6 months |  |  |  | 6-11 months |  |  |  | 12-23 months |  |  |  | 24-59 months |  |  |  |
| --- | --- | --- | --- | --- | --- | --- | --- | --- | --- | --- | --- | --- | --- | --- | --- | --- |
| Indicator | N | Death | Risk<sup>1</sup> | 95% CI | N | Death | Risk<sup>1</sup> | 95% CI | N | Death | Risk<sup>1</sup> | 95% CI | N | Death | Risk<sup>1</sup> | 95% CI |
| BF & WAZ ≥-2 | 78,448 | 72 | 0.4 | 0.2, 0.8 | 70,186 | 99 | 0.4 | 0.2, 0.8 | 50,120 | 37 | 0.4 | 0.2, 0.8 | 1,608 | 1 | 0.4 | 0.2, 0.9 |
| No BF & WAZ ≥-2 | 706 | 3 | 1.4 | 0.5, 4.3 | 2,110 | 10 | 1.0 | 0.4, 2.5 | 12,040 | 9 | 0.7 | 0.3, 1.8 | 8,897 | 1 | 0.5 | 0.2, 1.7 |
| BF & WAZ <-2 | 9,564 | 58 | 2.0 | 0.9, 4.6 | 17,841 | 76 | 1.5 | 0.7, 3.3 | 18,938 | 44 | 1.1 | 0.5, 2.5 | 981 | 4 | 0.8 | 0.3, 2.1 |
| No BF & WAZ <-2 | 192 | 5 | 5.3 | 1.5, 18.4 | 648 | 4 | 3.7 | 1.5, 9.2 | 5,466 | 7 | 2.6 | 1.0, 6.3 | 5,454 | 4 | 1.8 | 0.5, 5.9 |
| NBW & WAZ ≥-2 | 75,955 | 241 | 0.9 | 0.4, 2.0 | 57,350 | 94 | 0.6 | 0.2, 1.3 | 35,137 | 34 | 0.4 | 0.1, 0.8 | 14,458 | 1 | 0.2 | 0.1, 0.6 |
| LBW & WAZ ≥-2 | 4,962 | 31 | 1.6 | 0.7, 4.0 | 4,055 | 10 | 0.8 | 0.3, 2.0 | 2,524 | 3 | 0.4 | 0.1, 1.3 | 1,159 | 0 | 0.2 | 0.0, 0.9 |
| NBW & WAZ <-2 | 5,032 | 30 | 2.3 | 0.9, 5.5 | 8,697 | 50 | 1.6 | 0.7, 3.8 | 7,419 | 15 | 1.1 | 0.5, 2.8 | 4,681 | 1 | 0.8 | 0.3, 2.3 |
| LBW & WAZ <-2 | 5,808 | 140 | 6.6 | 2.8, 15.3 | 3,689 | 19 | 2.5 | 1.0, 6.1 | 2,693 | 10 | 1.0 | 0.4, 2.7 | 1,352 | 0 | 0.4 | 0.1, 1.3 |
| Term & WAZ ≥-2 | 73,293 | 245 | 1.7 | 1.0, 3.1 | 55,343 | 94 | 1.0 | 0.6, 1.9 | 32,746 | 33 | 0.6 | 0.3, 1.1 | 9,632 | 1 | 0.4 | 0.2, 0.7 |
| PTB & WAZ ≥-2 | 9,723 | 31 | 2.4 | 1.2, 4.7 | 8,232 | 19 | 1.7 | 0.9, 3.2 | 4,622 | 7 | 1.2 | 0.5, 2.6 | 784 | 0 | 0.8 | 0.3, 2.3 |
| Term & WAZ <-2 | 5,598 | 75 | 6.2 | 3.3, 11.4 | 7,662 | 54 | 3.6 | 2.0, 6.6 | 5,305 | 20 | 2.1 | 1.1, 4.1 | 1,192 | 1 | 1.2 | 0.6, 2.7 |
| PTB & WAZ <-2 | 2,496 | 97 | 22.3 | 12.2, 40.7 | 1,802 | 18 | 8.9 | 4.7, 17.0 | 1,126 | 9 | 3.6 | 1.6, 8.1 | 163 | 0 | 1.4 | 0.5, 4.1 |
| BF & WAZ ≥-3 | 85,076 | 87 | 0.4 | 0.2, 1.0 | 82,904 | 140 | 0.5 | 0.2, 1.0 | 63,805 | 60 | 0.5 | 0.2, 1.1 | 2,331 | 4 | 0.5 | 0.2, 1.2 |
| No BF & WAZ ≥-3 | 796 | 3 | 1.3 | 0.4, 4.0 | 2,537 | 11 | 1.1 | 0.5, 2.6 | 15,948 | 14 | 0.9 | 0.4, 2.1 | 12,969 | 2 | 0.7 | 0.3, 2.2 |
| BF & WAZ <-3 | 2,936 | 43 | 4.8 | 2.1, 10.8 | 5,123 | 35 | 2.8 | 1.3, 6.0 | 5,253 | 21 | 1.6 | 0.7, 3.8 | 258 | 1 | 0.9 | 0.3, 2.6 |
| No BF & WAZ <-3 | 102 | 5 | 13.2 | 3.7, 45.7 | 221 | 3 | 7.8 | 3.0, 19.9 | 1,558 | 2 | 4.6 | 1.6, 12.9 | 1,382 | 3 | 2.7 | 0.6, 11.7 |
| NBW & WAZ ≥-3 | 79,918 | 254 | 0.9 | 0.4, 2.1 | 64,021 | 122 | 0.6 | 0.3, 1.4 | 41,024 | 41 | 0.4 | 0.2, 1.0 | 18,321 | 2 | 0.3 | 0.1, 0.7 |
| LBW & WAZ ≥-3 | 8,422 | 69 | 2.1 | 0.9, 5.0 | 6,116 | 14 | 0.9 | 0.3, 2.1 | 4,114 | 4 | 0.3 | 0.1, 1.0 | 1,989 | 0 | 0.1 | 0.0, 0.5 |
| NBW & WAZ <-3 | 1,069 | 17 | 5.0 | 2.0, 12.8 | 2,026 | 22 | 3.6 | 1.5, 8.6 | 1,532 | 8 | 2.6 | 0.9, 6.9 | 818 | 0 | 1.8 | 0.5, 6.3 |
| LBW & WAZ <-3 | 2,348 | 102 | 13.5 | 5.8, 31.3 | 1,628 | 15 | 5.1 | 2.1, 12.3 | 1,103 | 8 | 1.9 | 0.7, 5.5 | 522 | 0 | 0.7 | 0.2, 2.7 |
| Term & WAZ ≥-3 | 77,551 | 280 | 1.9 | 1.1, 3.4 | 61,205 | 120 | 1.2 | 0.7, 2.1 | 37,010 | 42 | 0.7 | 0.4, 1.3 | 10,644 | 2 | 0.4 | 0.2, 0.8 |
| PTB & WAZ ≥-3 | 11,066 | 49 | 3.2 | 1.7, 6.0 | 9,423 | 25 | 2.1 | 1.1, 3.8 | 5,441 | 10 | 1.3 | 0.6, 2.8 | 913 | 0 | 0.8 | 0.3, 2.2 |
| Term & WAZ <-3 | 1,340 | 40 | 13.4 | 7.0, 25.3 | 1,800 | 28 | 8.5 | 4.6, 15.7 | 1,041 | 11 | 5.4 | 2.5, 11.3 | 180 | 0 | 3.4 | 1.3, 9.0 |
| PTB & WAZ <-3 | 1,153 | 79 | 40.1 | 22.0, 72.1 | 611 | 12 | 17.9 | 9.1, 34.9 | 307 | 6 | 7.9 | 3.1, 20.2 | 34 | 0 | 3.5 | 0.9, 12.6 |
| BF & HAZ ≥-2 | 73,307 | 71 | 0.4 | 0.2, 1.0 | 68,367 | 113 | 0.4 | 0.2, 0.9 | 41,912 | 38 | 0.4 | 0.2, 1.0 | 1,310 | 2 | 0.4 | 0.2, 1.0 |
| No BF & HAZ ≥-2 | 632 | 3 | 1.6 | 0.5, 4.8 | 1,983 | 10 | 1.3 | 0.6, 3.1 | 8,186 | 11 | 1.1 | 0.5, 2.6 | 6,453 | 1 | 0.9 | 0.3, 2.9 |
| BF & HAZ <-2 | 13,276 | 58 | 1.6 | 0.7, 3.4 | 19,417 | 62 | 0.8 | 0.5, 2.4 | 27,001 | 42 | 0.8 | 0.4, 1.8 | 1,279 | 3 | 0.6 | 0.2, 1.4 |
| No BF & HAZ <-2 | 261 | 5 | 3.3 | 1.0, 11.2 | 776 | 4 | 1.3 | 0.8, 5.1 | 9,123 | 4 | 1.3 | 0.5, 3.2 | 7,783 | 4 | 0.8 | 0.2, 2.7 |
| NBW & HAZ ≥-2 | 71,054 | 221 | 0.9 | 0.4, 2.0 | 55,216 | 103 | 0.6 | 0.3, 1.4 | 29,039 | 32 | 0.4 | 0.2, 1.0 | 10,737 | 0 | 0.3 | 0.1, 0.8 |
| LBW & HAZ ≥-2 | 5,116 | 36 | 1.7 | 0.7, 4.2 | 4,002 | 14 | 1.2 | 0.5, 2.9 | 1,903 | 5 | 0.8 | 0.3, 2.3 | 811 | 0 | 0.9 | 0.1, 2.1 |
| NBW & HAZ <-2 | 8,591 | 45 | 1.9 | 0.8, 4.6 | 10,650 | 41 | 1.2 | 0.5, 2.7 | 13,251 | 17 | 0.7 | 0.3, 1.7 | 8,269 | 2 | 0.4 | 0.2, 1.1 |
| LBW & HAZ <-2 | 5,540 | 133 | 6.7 | 2.9, 15.6 | 3,708 | 15 | 2.0 | 0.8, 4.8 | 3,299 | 8 | 0.6 | 0.2, 1.7 | 1,689 | 0 | 0.2 | 0.0, 0.6 |
| Term & HAZ ≥-2 | 68,724 | 240 | 1.8 | 0.9, 3.4 | 52,045 | 101 | 1.1 | 0.6, 2.1 | 25,958 | 28 | 0.7 | 0.4, 1.4 | 7,046 | 0 | 0.4 | 0.2, 0.9 |
| PTB & HAZ ≥-2 | 8,451 | 29 | 2.5 | 1.2, 5.1 | 7,188 | 18 | 2.3 | 1.1, 4.5 | 3,191 | 9 | 2.0 | 0.9, 4.7 | 467 | 0 | 1.8 | 0.6, 5.4 |
| Term & HAZ <-2 | 9,234 | 75 | 4.4 | 2.3, 8.5 | 10,841 | 47 | 2.3 | 1.2, 4.5 | 11,886 | 24 | 1.2 | 0.6, 2.5 | 3,653 | 2 | 0.7 | 0.3, 1.5 |
| PTB & HAZ <-2 | 3,567 | 97 | 17.4 | 9.0, 33.2 | 2,801 | 19 | 5.3 | 2.7, 10.7 | 2,533 | 7 | 1.6 | 0.7, 3.9 | 457 | 0 | 0.5 | 0.2, 1.5 |
| BF & WHZ ≥-2 | 81,063 | 100 | 0.5 | 0.2, 1.1 | 78,155 | 123 | 0.5 | 0.2, 1.0 | 60,701 | 58 | 0.5 | 0.2, 1.0 | 2,311 | 3 | 0.4 | 0.2, 1.0 |
| No BF & WHZ ≥-2 | 789 | 6 | 2.2 | 0.8, 6.0 | 2,439 | 12 | 1.4 | 0.6, 3.1 | 15,545 | 11 | 0.9 | 0.4, 2.0 | 12,899 | 3 | 0.6 | 0.2, 1.6 |
| BF & WHZ <-2 | 5,194 | 25 | 1-6 | 0-7, 3-6 | 9,529 | 52 | 1-5 | 0-7, 3-1 | 8,182 | 22 | 1-4 | 0-6, 3-1 | 277 | 2 | 1-3 | 0-5, 3-5 |
| No BF & WHZ <-2 | 101 | 2 | 3-0 | 0-6, 15-3 | 317 | 2 | 3-3 | 1-1, 9-8 | 1,760 | 4 | 3-7 | 1-4, 10-1 | 1,325 | 2 | 4-2 | 1-0, 18-3 |
| NBW & WHZ ≥-2 | 74,596 | 241 | 1-0 | 0-5, 2-2 | 60,139 | 109 | 0-7 | 0-3, 1-4 | 38,869 | 36 | 0-4 | 0-2, 0-9 | 17,957 | 2 | 0-3 | 0-1, 0-6 |
| LBW & WHZ ≥-2 | 7,719 | 65 | 2-3 | 1-0, 5-2 | 5,826 | 15 | 0-9 | 0-4, 2-1 | 3,931 | 4 | 0-3 | 0-1, 1-0 | 2,086 | 0 | 0-1 | 0-0, 0-5 |
| NBW & WHZ <-2 | 4,447 | 18 | 1-4 | 0-6, 3-3 | 5,634 | 35 | 1-7 | 0-8, 3-8 | 3,380 | 13 | 2-1 | 0-9, 4-9 | 1,037 | 0 | 2-5 | 0-9, 7-0 |
| LBW & WHZ <-2 | 1,896 | 32 | 4-0 | 1-7, 9-3 | 1,879 | 14 | 3-3 | 1-4, 7-4 | 1,269 | 9 | 2-7 | 1-0, 7-2 | 414 | 0 | 2-2 | 0-6, 7-7 |
| Term & WHZ ≥-2 | 73,105 | 268 | 2-0 | 1-2, 3-4 | 57,925 | 110 | 1-2 | 0-7, 2-0 | 35,400 | 37 | 0-7 | 0-4, 1-2 | 10,328 | 2 | 0-4 0-8 | 0-2, 0-7 |
| PTB & WHZ ≥-2 | 10,801 | 49 | 3-4 | 1-9, 6-0 | 9,121 | 27 | 2-1 | 1-2, 3-7 | 5,272 | 9 | 1-3 | 0-6, 2-6 | 891 | 0 | 3-8 | 0-3, 1-9 |
| Term & WHZ <-2 | 4,249 | 35 | 3-9 | 2-2, 7-1 | 4,878 | 38 | 3-9 | 2-2, 6-8 | 2,413 | 15 | 3-8 | 2-0, 7-3 | 362 | 0 | 10-3 | 1-6, 8-7 |
| PTB & WHZ <-2 | 729 | 14 | 9-7 | 4-7, 20-0 | 851 | 10 | 9-9 | 5-3, 18-6 | 441 | 7 | 10-1 | 4-3, 23-9 | 32 | 0 |  | 2-9, 35-7 |
| BF & MUAC ≥125 | 34,445 | 42 | 1-0 | 0-6, 1-9 | 63,418 | 110 | 1-0 | 0-6, 1-8 | 58,937 | 59 | 1-1 | 0-6, 1-9 | 936 | 4 | 1-1 | 0-5, 2-1 |
| No BF & MUAC ≥125 | 452 | 3 | 3-2 | 1-1, 9-1 | 1,316 | 11 | 2-8 | 1-4, 5-6 | 10,574 | 12 | 2-4 | 1-2, 4-8 | 3,468 | 2 | 2-1 | 0-7, 5-9 |
| BF & MUAC <125 | 13,304 | 35 | 2-1 | 1-1, 3-9 | 8,251 | 47 | 2-8 | 1-6, 4-9 | 5,104 | 21 | 3-7 | 2-0, 7-1 | 64 | 1 | 5-0 | 2-2, 11-4 |
| No BF & MUAC <125 | 141 | 3 | 5-4 | 1-2, 23-1 | 94 | 1 | 6-6 | 2-6, 16-6 | 1,108 | 2 | 8-0 | 2-9, 22-0 | 264 | 2 | 9-7 | 1-9, 48-2 |
| NBW & MUAC ≥125 | 38,552 | 99 | 1-4 | 0-8, 2-6 | 48,363 | 108 | 1-1 | 0-6, 2-0 | 32,485 | 41 | 0-9 | 0-3, 1-7 | 8,944 | 2 | 0-7 | 0-3, 1-4 |
| NBW & MUAC ≥125 | 3,225 | 16 | 2-2 | 1-1, 4-7 | 4,865 | 16 | 1-7 | 0-9, 3-4 | 3,107 | 8 | 1-4 | 0-3, 3-1 | 745 | 0 | 1-1 | 0-3, 3-2 |
| LBW & MUAC <125 | 22,849 | 153 | 2-8 | 1-5, 5-1 | 3,792 | 22 | 3-2 | 1-7, 6-2 | 1,291 | 8 | 3-7 | 1-7, 8-3 | 47 | 0 | 4-3 | 1-5, 12-0 |
| LBW & MUAC <125 | 5,611 | 142 | 11-5 | 6-3, 21-1 | 1,425 | 10 | 5-5 | 2-7, 11-2 | 560 | 5 | 2-6 | 0-9, 7-2 | 24 | 0 | 1-2 | 0-3, 5-0 |
| Term & MUAC ≥125 | 35,504 | 98 | 1-5 | 0-7, 3-1 | 44,378 | 100 | 1-1 | 0-5, 2-3 | 30,080 | 36 | 0-8 | 0-4, 1-8 | 9,456 | 2 | 0-6 | 0-3, 1-4 |
| PTB & MUAC ≥125 | 3,818 | 17 | 2-9 | 1-2, 6-5 | 5,722 | 19 | 2-5 | 1-2, 5-4 | 3,704 | 11 | 2-2 | 0-9, 5-1 | 559 | 0 | 1-9 | 0-6, 5-6 |
| Term & MUAC <125 | 21,433 | 182 | 3-3 | 1-6, 6-7 | 3,664 | 19 | 3-2 | 1-5, 6-9 | 1,283 | 10 | 3-2 | 1-3, 7-8 | 59 | 0 | 3-1 | 1-0, 9-4 |
| PTB & MUAC <125 | 3,578 | 86 | 13-9 | 6-7, 28-6 | 689 | 7 | 8-8 | 3-8, 20-4 | 264 | 4 | 5-6 | 1-7, 18-1 | 12 | 0 | - | - |

<sup>1</sup> Risk = predicted absolute risk of dying within 1 month per 1,000 child months modelled using generalized linear mixed effects models accounting for repeated measures in each child, clustering within studies, and time at risk
Abbreviations: BF = breastfeeding; HAZ = height-for-age Z-score; LBW= low birthweight (<2500g); MUAC = mid-upper arm circumference; NBW = normal birthweight (≥2500g); PTB = preterm birth (<37 weeks of gestation at birth); WAZ = weight-for-age Z-score; WHZ= weight-for-height Z-score

### Mortality risks in the anthropometry-selected (A-S) and illness-selected (I-S) populations

Although children in the A-S population were older than those in the GP population, mortality was similar in both groups (2·5% in both). However, in the I-S population, mortality was significantly higher (11%) and did not decrease as clearly with increasing age (Supplementary Table S6).

Anthropometric deficits in the A-S and I-S populations tended to be more severe, but these deficits did not identify mortality risk as clearly as in GP studies. (Tables S6 and S7) There was only one A-S study with information on birthweight and LBW did not increase the risk of death. In I-S studies, there was an increased mortality risk in LBW and PTB (although estimates for PTB were imprecise due to low number of preterm children (Tables S8 and S9).

Among infants less than six months of age in I-S studies, the mortality risk of those with an IMCI danger sign was six times higher compared to those without. The absolute mortality risk decreased with age but remained higher than in children without danger signs in each age group. The risk increased if there was more than one danger sign present – though confidence intervals were overlapping. There were too few children in A-S studies with multiple dangers signs to enable analyses.

In both the A-S and the I-S child populations, the mortality risk of not being breastfed added to the risks associated with anthropometric deficits for all measures, although sample sizes were limited with overlapping confidence intervals (Tables 3 and 4).

**Table 3:** Absolute risk of death per 1000 child months by age group and by combinations of exposures in Anthropometry-Selected (A-S) cohorts, i.e., enrolment on the basis of anthropometric deficit.

|  | <6 months |  |  |  | 6-11 months |  |  |  | 12-23 months |  |  |  | 24-59 months |  |  |  |
| --- | --- | --- | --- | --- | --- | --- | --- | --- | --- | --- | --- | --- | --- | --- | --- | --- |
| Indicator | N | Deaths | Risk<sup>2</sup> | 95% CI | N | Deaths | Risk<sup>2</sup> | 95% CI | N | Deaths | Risk<sup>2</sup> | 95% CI | N | Deaths | Risk<sup>2</sup> | 95% CI |
| BF & WAZ ≥-2 | 4,123 | 0 | 1.3 | 0.2, 8.9 | 2,439 | 8 | 1.6 | 0.3, 7.4 | 1,944 | 3 | 2.0 | 0.4, 9.5 | 113 | 1 | 2.5 | 0.3, 18.2 |
| No BF & WAZ ≥-2 | 40 | 0 | 0.8 | 0.0, 20.4 | 142 | 0 | 1.3 | 0.1, 13.7 | 983 | 4 | 2.3 | 0.4, 11.5 | 1,314 | 7 | 3.8 | 0.8, 17.4 |
| BF & WAZ <-2 | 3,706 | 16 | 13.3 | 3.3, 51.7 | 6,997 | 79 | 9.6 | 2.5, 36.3 | 9,283 | 74 | 7.0 | 1.8, 26.8 | 1,206 | 10 | 5.0 | 1.2, 20.7 |
| No BF & WAZ <-2 | 52 | 1 | 33.9 | 7.7, 136 | 390 | 15 | 19.4 | 4.8, 74.8 | 3,129 | 48 | 11.0 | 2.8, 41.8 | 5,041 | 49 | 6.2 | 1.6, 24.3 |
| NBW & WAZ ≥-2 | 1 | 0 | .. | .. | 570 | 5 | 7.5 | 3.2, 17.6 | 507 | 2 | 3.6 | 1.0, 13.4 | 64 | 0 | 1.8 | 0.1, 25.7 |
| LBW & WAZ ≥-2 | 0 | 0 | .. | .. | 10 | 0 | .. | .. | 17 | 0 | .. | .. | 4 | 0 | .. | .. |
| NBW & WAZ <-2 | 0 | 0 | .. | .. | 1,564 | 15 | 10.1 | 6.4, 16.2 | 2,141 | 19 | 8.5 | 5.9, 12.2 | 528 | 3 | 7.1 | 3.4, 15.0 |
| LBW & WAZ <-2 | 0 | 0 | .. | .. | 90 | 1 | 7.8 | 1.1, 54.7 | 117 | 1 | 12.5 | 4.0, 38.4 | 37 | 1 | 20.1 | 2.7, 134 |
| Term & WAZ ≥-2 | 0 | 0 | .. | .. | 0 | 0 | .. | .. | 0 | 0 | .. | .. | 0 | 0 | .. | .. |
| PTB & WAZ ≥-2 | 0 | 0 | .. | .. | 0 | 0 | .. | .. | 0 | 0 | .. | .. | 0 | 0 | .. | .. |
| Term & WAZ <-2 | 0 | 0 | .. | .. | 0 | 0 | .. | .. | 0 | 0 | .. | .. | 0 | 0 | .. | .. |
| PTB & WAZ <-2 | 0 | 0 | .. | .. | 0 | 0 | .. | .. | 0 | 0 | .. | .. | 0 | 0 | .. | .. |
| BF & WAZ ≥-3 | 6,814 | 3 | 2.7 | 0.6, 11.3 | 5,911 | 23 | 2.8 | 0.8, 10.3 | 6,122 | 17 | 3.0 | 0.8, 11.0 | 480 | 1 | 3.1 | 0.7, 13.7 |
| No BF & WAZ ≥-3 | 65 | 0 | 6.0 | 1.0, 36.4 | 266 | 3 | 5.8 | 1.3, 25.6 | 2,259 | 14 | 5.6 | 1.5, 20.5 | 3,202 | 21 | 5.4 | 1.4, 19.9 |
| BF & WAZ <-3 | 1,015 | 13 | 22.9 | 6.3, 79.8 | 3,525 | 64 | 15.4 | 4.4, 52.1 | 5,105 | 60 | 10.3 | 2.9, 35.7 | 839 | 10 | 6.9 | 1.8, 25.8 |
| No BF & WAZ <-3 | 27 | 1 | 48.4 | 11.8, 177 | 266 | 12 | 26.4 | 7.2, 92.5 | 1,853 | 38 | 14.2 | 4.0, 49.0 | 3,153 | 35 | 7.6 | 2.1, 27.2 |
| NBW & WAZ ≥-3 | 1 | 0 | .. | .. | 1,590 | 12 | 7.7 | 4.6, 13.1 | 1,661 | 12 | 6.1 | 3.6, 10.4 | 219 | 0 | 4.9 | 1.6, 14.7 |
| LBW & WAZ ≥-3 | 0 | 0 | .. | .. | 34 | 0 | 6.9 | 0.2, 174 | 60 | 1 | 7.7 | 1.1, 52.7 | 25 | 0 | 8.5 | 0.3, 209 |
| NBW & WAZ <-3 | 0 | 0 | .. | .. | 544 | 8 | 14.4 | 7.4, 27.7 | 987 | 9 | 10.2 | 6.4, 16.3 | 373 | 3 | 7.2 | 2.8, 18.4 |
| LBW & WAZ <-3 | 0 | 0 | .. | .. | 66 | 1 | 6.5 | 0.5, 73.4 | 74 | 0 | 14.8 | 3.7, 57.9 | 16 | 1 | .. | .. |
| Term & WAZ ≥-3 | 0 | 0 | .. | .. | 0 | 0 | .. | .. | 0 | 0 | .. | .. | 0 | 0 | .. | .. |
| PTB & WAZ ≥-3 | 0 | 0 | .. | .. | 0 | 0 | .. | .. | 0 | 0 | .. | .. | 0 | 0 | .. | .. |
| Term & WAZ <-3 | 0 | 0 | .. | .. | 0 | 0 | .. | .. | 0 | 0 | .. | .. | 0 | 0 | .. | .. |
| PTB & WAZ <-3 | 0 | 0 | .. | .. | 0 | 0 | .. | .. | 0 | 0 | .. | .. | 0 | 0 | .. | .. |
| BF & HAZ ≥-2 | 4,099 | 2 | 5.1 | 1.0, 24.8 | 3,775 | 19 | 3.4 | 0.8, 14.1 | 2,357 | 5 | 2.3 | 0.5, 10.8 | 158 | 0 | 1.6 | 0.2, 10.5 |
| No BF & HAZ ≥-2 | 41 | 0 | 2.6 | 0.2, 32.8 | 193 | 1 | 3.2 | 0.5, 21.4 | 823 | 5 | 3.9 | 0.9, 17.7 | 1,115 | 6 | 4.8 | 1.0, 22.4 |
| BF & HAZ <-2 | 3,643 | 14 | 14.8 | 3.6, 58.4 | 5,646 | 68 | 10.1 | 2.6, 38.8 | 8,865 | 72 | 6.9 | 1.7, 26.9 | 1,161 | 11 | 4.7 | 1.1, 19.6 |
| No BF & HAZ <-2 | 50 | 1 | 36.3 | 8.1, 148 | 339 | 14 | 19.6 | 4.8, 76.9 | 3,288 | 47 | 10.5 | 2.7, 40.5 | 5,237 | 50 | 5.6 | 1.4, 22.2 |
| NBW & HAZ ≥-2 | 1 | 0 | .. | .. | 765 | 6 | 7.4 | 3.4, 16.1 | 481 | 3 | 6.0 | 2.1, 17.1 | 52 | 0 | 4.9 | 0.6, 41.1 |
| LBW & HAZ ≥-2 | 0 | 0 | .. | .. | 10 | 0 | .. | .. | 15 | 0 | .. | .. | 13 | 0 | .. | .. |
| NBW & HAZ <-2 | 0 | 0 | .. | .. | 1,367 | 14 | 10.4 | 6.4, 16.9 | 2,162 | 18 | 8.1 | 5.6, 11.7 | 539 | 3 | 6.3 | 2.9, 13.5 |
| LBW & HAZ <-2 | 0 | 0 | .. | .. | 90 | 1 | 7.3 | 1.0, 52.7 | 119 | 1 | 12.9 | 4.1, 39.6 | 28 | 1 | 22.8 | 3.0, 153 |
| Term & HAZ ≥-2 | 0 | 0 | .. | .. | 0 | 0 | .. | .. | 0 | 0 | .. | .. | 0 | 0 | .. | .. |
| PTB & HAZ ≥-2 | 0 | 0 | .. | .. | 0 | 0 | .. | .. | 0 | 0 | .. | .. | 0 | 0 | .. | .. |
| Term & HAZ <-2 | 0 | 0 | .. | .. | 0 | 0 | .. | .. | 0 | 0 | .. | .. | 0 | 0 | .. | .. |
| PTB & HAZ <-2 | 0 | 0 | .. | .. | 0 | 0 | .. | .. | 0 | 0 | .. | .. | 0 | 0 | .. | .. |
| BF & WHZ ≥-2 | 6,966 | 8 | 3.8 | 0.9, 15.9 | 5,381 | 22 | 3.5 | 0.9, 13.1 | 5,917 | 15 | 3.2 | 0.8, 12.4 | 601 | 4 | 3.0 | 0.7, 13.4 |
| No BF & WHZ ≥-2 | 75 | 0 | 5.9 | 0.9, 37.0 | 259 | 2 | 5.5 | 1.2, 25.3 | 2,457 | 16 | 5.1 | 1.3, 19.6 | 3,854 | 24 | 4.7 | 1.2, 18.2 |
| BF & WHZ <-2 | 771 | 8 | 17.9 | 4.6, 66.4 | 4,038 | 65 | 13.1 | 3.6, 46.9 | 5,305 | 62 | 9.6 | 2.6, 35.0 | 717 | 7 | 7.0 | 1.8, 27.6 |
| No BF & WHZ <-2 | 16 | 1 | .. | .. | 273 | 13 | 24.2 | 6.2, 89.1 | 1,652 | 36 | 14.7 | 4.0, 52.7 | 2,497 | 32 | 8.8 | 2.3, 33.0 |
| NBW & WHZ ≥-2 | 1 | 0 | .. | .. | 1,645 | 15 | 8.5 | 5.2, 13.9 | 1,754 | 11 | 6.6 | 4.1, 10.5 | 338 | 2 | 5.1 | 1.9, 13.3 |
| LBW & WHZ ≥-2 | 0 | 0 | .. | .. | 81 | 1 | 12.6 | 2.1, 73.1 | 91 | 1 | 7.7 | 1.3, 45.0 | 20 | 0 | 4.6 | 0.1, 162 |
| NBW & WHZ <-2 | 0 | 0 | .. | .. | 486 | 5 | 12.1 | 5.7, 25.5 | 888 | 10 | 10.0 | 5.9, 16.8 | 253 | 1 | 8.2 | 2.8, 23.8 |
| LBW & WHZ <-2 | 0 | 0 | .. | .. | 19 | 0 | .. | .. | 43 | 0 | 0.0 | 0, 1000 | 21 | 1 | 57.9 | 7.7, 327 |
| Term & WHZ ≥-2 | 0 | 0 | .. | .. | 0 | 0 | .. | .. | 0 | 0 | .. | .. | 0 | 0 | .. | .. |
| PTB & WHZ ≥-2 | 0 | 0 | .. | .. | 0 | 0 | .. | .. | 0 | 0 | .. | .. | 0 | 0 | .. | .. |
| Term & WHZ <-2 | 0 | 0 | .. | .. | 0 | 0 | .. | .. | 0 | 0 | .. | .. | 0 | 0 | .. | .. |
| PTB & WHZ <-2 | 0 | 0 | .. | .. | 0 | 0 | .. | .. | 0 | 0 | .. | .. | 0 | 0 | .. | .. |
| BF & MUAC ≥125 | 3,226 | 1 | 2.5 | 0.4, 15.2 | 2,201 | 6 | 2.1 | 0.4, 9.9 | 3,903 | 8 | 1.8 | 0.4, 8.3 | 602 | 1 | 1.5 | 0.2, 9.0 |
| No BF & MUAC ≥125 | 42 | 0 | 7.9 | 1.0, 60.8 | 157 | 2 | 5.9 | 1.0, 32.8 | 1,737 | 11 | 4.5 | 1.0, 20.1 | 4,072 | 21 | 3.4 | 0.8, 15.0 |
| BF & MUAC <125 | 4,573 | 14 | 10.3 | 2.4, 44.2 | 7,207 | 81 | 9.0 | 2.1, 37.3 | 7,301 | 69 | 7.9 | 1.9, 33.1 | 708 | 10 | 6.9 | 1.5, 30.9 |
| No BF & MUAC <125 | 49 | 1 | 21.8 | 4.4, 100 | 372 | 13 | 15.8 | 3.6, 67.8 | 2,368 | 41 | 11.5 | 2.7, 47.7 | 2,264 | 35 | 8.3 | 1.9, 35.6 |
| NBW & MUAC ≥125 | 0 | 0 | .. | .. | 438 | 2 | 4.9 | 1.8, 13.2 | 949 | 8 | 5.9 | 3.3, 10.8 | 279 | 1 | 7.2 | 2.3, 22.1 |
| NBW & MUAC <125 | 0 | 0 | .. | .. | 15 | 0 | .. | .. | 47 | 0 | 0.0 | 0.0, 1000 | 18 | 0 | .. | .. |
| LBW & MUAC <125 | 1 | 0 | .. | .. | 1693 | 18 | 11.2 | 7.1, 17.5 | 1,700 | 13 | 8.4 | 5.4, 13.1 | 313 | 2 | 6.4 | 2.5, 15.8 |
| LBW & MUAC <125 | 0 | 0 | .. | .. | 85 | 1 | 8.8 | 1.2, 61.6 | 87 | 1 | 19.0 | 6.1, 58.0 | 23 | 1 | 40.5 | 5.5, 244 |
| Term & MUAC ≥125 | 0 | 0 | .. | .. | 0 | 0 | .. | .. | 0 | 0 | .. | .. | 0 | 0 | .. | .. |
| PTB & MUAC ≥125 | 0 | 0 | .. | .. | 0 | 0 | .. | .. | 0 | 0 | .. | .. | 0 | 0 | .. | .. |
| Term & MUAC <125 | 0 | 0 | .. | .. | 0 | 0 | .. | .. | 0 | 0 | .. | .. | 0 | 0 | .. | .. |
| PTB & MUAC <125 | 0 | 0 | .. | .. | 0 | 0 | .. | .. | 0 | 0 | .. | .. | 0 | 0 | .. | .. |

<sup>1</sup> Risk = predicted absolute risk of dying within 1 month per 1,000 child months modelled using generalized linear mixed effects models accounting for repeated measures in each child, clustering within studies, and time at risk
Abbreviations: BF = breastfeeding; HAZ = height-for-age Z-score; LBW= low birthweight (<2500g); MUAC = mid-upper arm circumference; NBW = normal birthweight (≥2500g); PTB = preterm birth (<37 weeks of gestation at birth); WAZ = weight-for-age Z-score; WHZ= weight-for-height Z-score

**Table 4:**
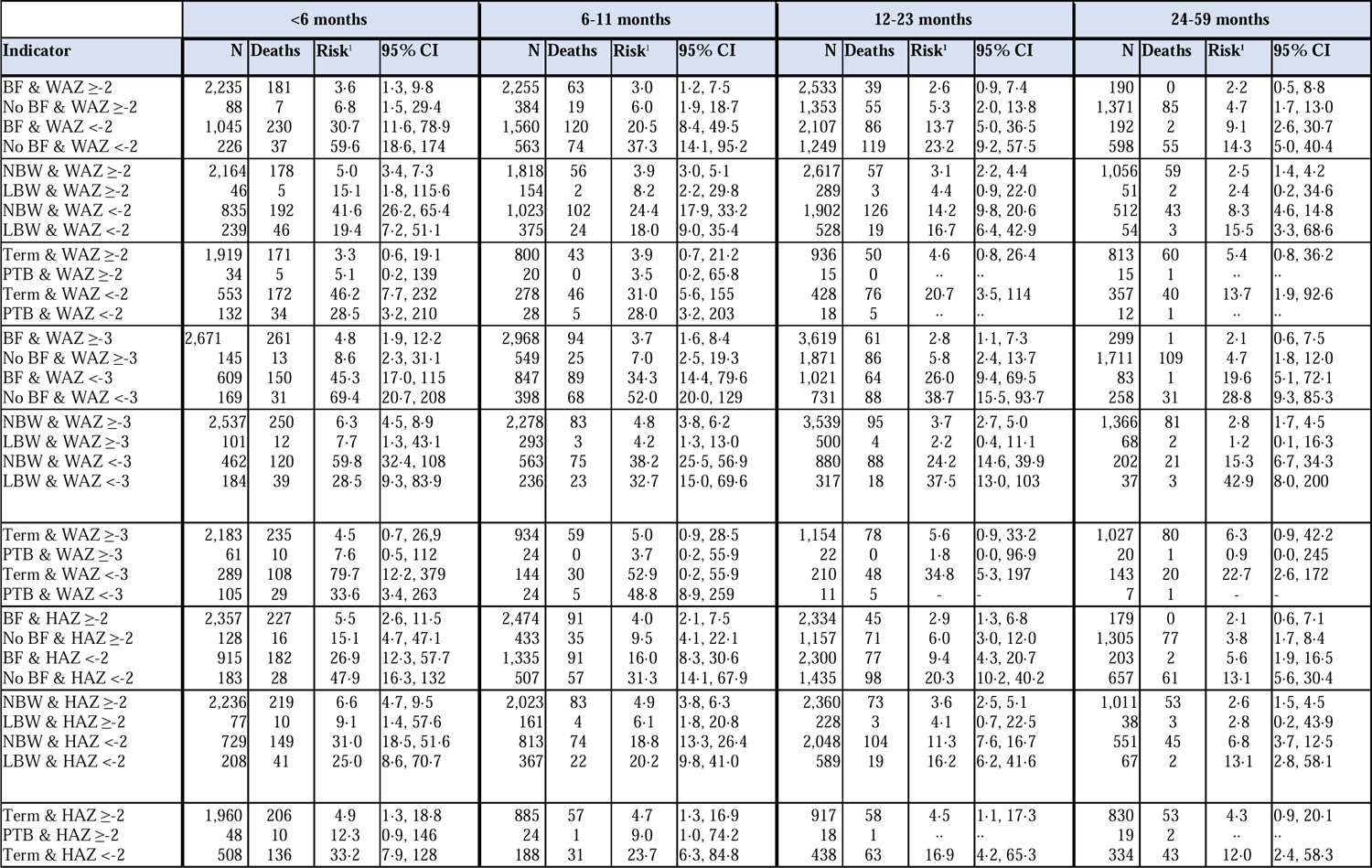
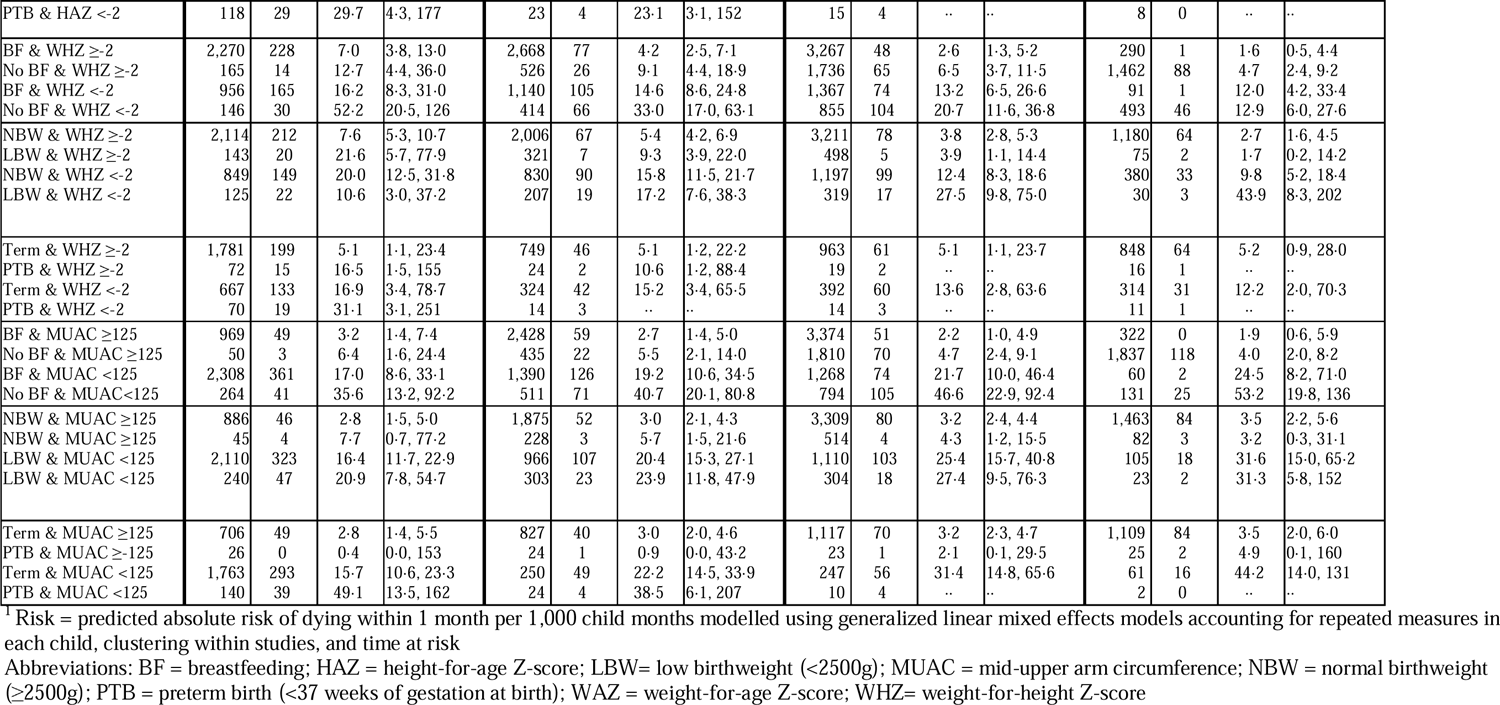
Absolute risk of death per 1000 child months by age group and by combinations of exposures in Illness-Selected (I-S) cohorts, i.e., enrolment on the basis of the presence of illness.

|  | <6 months |  |  |  | 6-11 months |  |  |  | 12-23 months |  |  |  | 24-59 months |  |  |  |
| --- | --- | --- | --- | --- | --- | --- | --- | --- | --- | --- | --- | --- | --- | --- | --- | --- |
| Indicator | N | Deaths | Risk<sup>1</sup> | 95% CI | N | Deaths | Risk<sup>1</sup> | 95% CI | N | Deaths | Risk<sup>1</sup> | 95% CI | N | Deaths | Risk<sup>1</sup> | 95% CI |
| BF & WAZ ≥-2 | 2,235 | 181 | 3.6 | 1.3, 9.8 | 2,255 | 63 | 3.0 | 1.2, 7.5 | 2,533 | 39 | 2.6 | 0.9, 7.4 | 190 | 0 | 2.2 | 0.5, 8.8 |
| No BF & WAZ ≥-2 | 88 | 7 | 6.8 | 1.5, 29.4 | 384 | 19 | 6.0 | 1.9, 18.7 | 1,353 | 55 | 5.3 | 2.0, 13.8 | 1,371 | 85 | 4.7 | 1.7, 13.0 |
| BF & WAZ <-2 | 1,045 | 230 | 30.7 | 11.6, 78.9 | 1,560 | 120 | 20.5 | 8.4, 49.5 | 2,107 | 86 | 13.7 | 5.0, 36.5 | 192 | 2 | 9.1 | 2.6, 30.7 |
| No BF & WAZ <-2 | 226 | 37 | 59.6 | 18.6, 174 | 563 | 74 | 37.3 | 14.1, 95.2 | 1,249 | 119 | 23.2 | 9.2, 57.5 | 598 | 55 | 14.3 | 5.0, 40.4 |
| NBW & WAZ ≥-2 | 2,164 | 178 | 5.0 | 3.4, 7.3 | 1,818 | 56 | 3.9 | 3.0, 5.1 | 2,617 | 57 | 3.1 | 2.2, 4.4 | 1,056 | 59 | 2.5 | 1.4, 4.2 |
| LBW & WAZ ≥-2 | 46 | 5 | 15.1 | 1.8, 115.6 | 154 | 2 | 8.2 | 2.2, 29.8 | 289 | 3 | 4.4 | 0.9, 22.0 | 51 | 2 | 2.4 | 0.2, 34.6 |
| NBW & WAZ <-2 | 835 | 192 | 41.6 | 26.2, 65.4 | 1,023 | 102 | 24.4 | 17.9, 33.2 | 1,902 | 126 | 14.2 | 9.8, 20.6 | 512 | 43 | 8.3 | 4.6, 14.8 |
| LBW & WAZ <-2 | 239 | 46 | 19.4 | 7.2, 51.1 | 375 | 24 | 18.0 | 9.0, 35.4 | 528 | 19 | 16.7 | 6.4, 42.9 | 54 | 3 | 15.5 | 3.3, 68.6 |
| Term & WAZ ≥-2 | 1,919 | 171 | 3.3 | 0.6, 19.1 | 800 | 43 | 3.9 | 0.7, 21.2 | 936 | 50 | 4.6 | 0.8, 26.4 | 813 | 60 | 5.4 | 0.8, 36.2 |
| PTB & WAZ ≥-2 | 34 | 5 | 5.1 | 0.2, 139 | 20 | 0 | 3.5 | 0.2, 65.8 | 15 | 0 | .. | .. | 15 | 1 | .. | .. |
| Term & WAZ <-2 | 553 | 172 | 46.2 | 7.7, 232 | 278 | 46 | 31.0 | 5.6, 155 | 428 | 76 | 20.7 | 3.5, 114 | 357 | 40 | 13.7 | 1.9, 92.6 |
| PTB & WAZ <-2 | 132 | 34 | 28.5 | 3.2, 210 | 28 | 5 | 28.0 | 3.2, 203 | 18 | 5 | .. | .. | 12 | 1 | .. | .. |
| BF & WAZ ≥-3 | 2,671 | 261 | 4.8 | 1.9, 12.2 | 2,968 | 94 | 3.7 | 1.6, 8.4 | 3,619 | 61 | 2.8 | 1.1, 7.3 | 299 | 1 | 2.1 | 0.6, 7.5 |
| No BF & WAZ ≥-3 | 145 | 13 | 8.6 | 2.3, 31.1 | 549 | 25 | 7.0 | 2.5, 19.3 | 1,871 | 86 | 5.8 | 2.4, 13.7 | 1,711 | 109 | 4.7 | 1.8, 12.0 |
| BF & WAZ <-3 | 609 | 150 | 45.3 | 17.0, 115 | 847 | 89 | 34.3 | 14.4, 79.6 | 1,021 | 64 | 26.0 | 9.4, 69.5 | 83 | 1 | 19.6 | 5.1, 72.1 |
| No BF & WAZ <-3 | 169 | 31 | 69.4 | 20.7, 208 | 398 | 68 | 52.0 | 20.0, 129 | 731 | 88 | 38.7 | 15.5, 93.7 | 258 | 31 | 28.8 | 9.3, 85.3 |
| NBW & WAZ ≥-3 | 2,537 | 250 | 6.3 | 4.5, 8.9 | 2,278 | 83 | 4.8 | 3.8, 6.2 | 3,539 | 95 | 3.7 | 2.7, 5.0 | 1,366 | 81 | 2.8 | 1.7, 4.5 |
| LBW & WAZ ≥-3 | 101 | 12 | 7.7 | 1.3, 43.1 | 293 | 3 | 4.2 | 1.3, 13.0 | 500 | 4 | 2.2 | 0.4, 11.1 | 68 | 2 | 1.2 | 0.1, 16.3 |
| NBW & WAZ <-3 | 462 | 120 | 59.8 | 32.4, 108 | 563 | 75 | 38.2 | 25.5, 56.9 | 880 | 88 | 24.2 | 14.6, 39.9 | 202 | 21 | 15.3 | 6.7, 34.3 |
| LBW & WAZ <-3 | 184 | 39 | 28.5 | 9.3, 83.9 | 236 | 23 | 32.7 | 15.0, 69.6 | 317 | 18 | 37.5 | 13.0, 103 | 37 | 3 | 42.9 | 8.0, 200 |
| Term & WAZ ≥-3 | 2,183 | 235 | 4.5 | 0.7, 26.9 | 934 | 59 | 5.0 | 0.9, 28.5 | 1,154 | 78 | 5.6 | 0.9, 33.2 | 1,027 | 80 | 6.3 | 0.9, 42.2 |
| PTB & WAZ ≥-3 | 61 | 10 | 7.6 | 0.5, 112 | 24 | 0 | 3.7 | 0.2, 55.9 | 22 | 0 | 1.8 | 0.0, 96.9 | 20 | 1 | 0.9 | 0.0, 245 |
| Term & WAZ <-3 | 289 | 108 | 79.7 | 12.2, 379 | 144 | 30 | 52.9 | 0.2, 55.9 | 210 | 48 | 34.8 | 5.3, 197 | 143 | 20 | 22.7 | 2.6, 172 |
| PTB & WAZ <-3 | 105 | 29 | 33.6 | 3.4, 263 | 24 | 5 | 48.8 | 8.9, 259 | 11 | 5 | - | - | 7 | 1 | - | - |
| BF & HAZ ≥-2 | 2,357 | 227 | 5.5 | 2.6, 11.5 | 2,474 | 91 | 4.0 | 2.1, 7.5 | 2,334 | 45 | 2.9 | 1.3, 6.8 | 179 | 0 | 2.1 | 0.6, 7.1 |
| No BF & HAZ ≥-2 | 128 | 16 | 15.1 | 4.7, 47.1 | 433 | 35 | 9.5 | 4.1, 22.1 | 1,157 | 71 | 6.0 | 3.0, 12.0 | 1,305 | 77 | 3.8 | 1.7, 8.4 |
| BF & HAZ <-2 | 915 | 182 | 26.9 | 12.3, 57.7 | 1,335 | 91 | 16.0 | 8.3, 30.6 | 2,300 | 77 | 9.4 | 4.3, 20.7 | 203 | 2 | 5.6 | 1.9, 16.5 |
| No BF & HAZ <-2 | 183 | 28 | 47.9 | 16.3, 132 | 507 | 57 | 31.3 | 14.1, 67.9 | 1,435 | 98 | 20.3 | 10.2, 40.2 | 657 | 61 | 13.1 | 5.6, 30.4 |
| NBW & HAZ ≥-2 | 2,236 | 219 | 6.6 | 4.7, 9.5 | 2,023 | 83 | 4.9 | 3.8, 6.3 | 2,360 | 73 | 3.6 | 2.5, 5.1 | 1,011 | 53 | 2.6 | 1.5, 4.5 |
| LBW & HAZ ≥-2 | 77 | 10 | 9.1 | 1.4, 57.6 | 161 | 4 | 6.1 | 1.8, 20.8 | 228 | 3 | 4.1 | 0.7, 22.5 | 38 | 3 | 2.8 | 0.2, 43.9 |
| NBW & HAZ <-2 | 729 | 149 | 31.0 | 18.5, 51.6 | 813 | 74 | 18.8 | 13.3, 26.4 | 2,048 | 104 | 11.3 | 7.6, 16.7 | 551 | 45 | 6.8 | 3.7, 12.5 |
| LBW & HAZ <-2 | 208 | 41 | 25.0 | 8.6, 70.7 | 367 | 22 | 20.2 | 9.8, 41.0 | 589 | 19 | 16.2 | 6.2, 41.6 | 67 | 2 | 13.1 | 2.8, 58.1 |
| Term & HAZ ≥-2 | 1,960 | 206 | 4.9 | 1.3, 18.8 | 885 | 57 | 4.7 | 1.3, 16.9 | 917 | 58 | 4.5 | 1.1, 17.3 | 830 | 53 | 4.3 | 0.9, 20.1 |
| PTB & HAZ ≥-2 | 48 | 10 | 12.3 | 0.9, 146 | 24 | 1 | 9.0 | 1.0, 74.2 | 18 | 1 | .. | .. | 19 | 2 | .. | .. |
| Term & HAZ <-2 | 508 | 136 | 33.2 | 7.9, 128 | 188 | 31 | 23.7 | 6.3, 84.8 | 438 | 63 | 16.9 | 4.2, 65.3 | 334 | 43 | 12.0 | 2.4, 58.3 |
| PTB & HAZ <-2 | 118 | 29 | 29.7 | 4.3, 177 | 23 | 4 | 23.1 | 3.1, 152 | 15 | 4 | .. | .. | 8 | 0 | .. | .. |
| BF & WHZ ≥-2 | 2,270 | 228 | 7.0 | 3.8, 13.0 | 2,668 | 77 | 4.2 | 2.5, 7.1 | 3,267 | 48 | 2.6 | 1.3, 5.2 | 290 | 1 | 1.6 | 0.5, 4.4 |
| No BF & WHZ ≥-2 | 165 | 14 | 12.7 | 4.4, 36.0 | 526 | 26 | 9.1 | 4.4, 18.9 | 1,736 | 65 | 6.5 | 3.7, 11.5 | 1,462 | 88 | 4.7 | 2.4, 9.2 |
| BF & WHZ <-2 | 956 | 165 | 16.2 | 8.3, 31.0 | 1,140 | 105 | 14.6 | 8.6, 24.8 | 1,367 | 74 | 13.2 | 6.5, 26.6 | 91 | 1 | 12.0 | 4.2, 33.4 |
| No BF & WHZ <-2 | 146 | 30 | 52.2 | 20.5, 126 | 414 | 66 | 33.0 | 17.0, 63.1 | 855 | 104 | 20.7 | 11.6, 36.8 | 493 | 46 | 12.9 | 6.0, 27.6 |
| NBW & WHZ ≥-2 | 2,114 | 212 | 7.6 | 5.3, 10.7 | 2,006 | 67 | 5.4 | 4.2, 6.9 | 3,211 | 78 | 3.8 | 2.8, 5.3 | 1,180 | 64 | 2.7 | 1.6, 4.5 |
| LBW & WHZ ≥-2 | 143 | 20 | 21.6 | 5.7, 77.9 | 321 | 7 | 9.3 | 3.9, 22.0 | 498 | 5 | 3.9 | 1.1, 14.4 | 75 | 2 | 1.7 | 0.2, 14.2 |
| NBW & WHZ <-2 | 849 | 149 | 20.0 | 12.5, 31.8 | 830 | 90 | 15.8 | 11.5, 21.7 | 1,197 | 99 | 12.4 | 8.3, 18.6 | 380 | 33 | 9.8 | 5.2, 18.4 |
| LBW & WHZ <-2 | 125 | 22 | 10.6 | 3.0, 37.2 | 207 | 19 | 17.2 | 7.6, 38.3 | 319 | 17 | 27.5 | 9.8, 75.0 | 30 | 3 | 43.9 | 8.3, 202 |
| Term & WHZ ≥-2 | 1,781 | 199 | 5.1 | 1.1, 23.4 | 749 | 46 | 5.1 | 1.2, 22.2 | 963 | 61 | 5.1 | 1.1, 23.7 | 848 | 64 | 5.2 | 0.9, 28.0 |
| PTB & WHZ ≥-2 | 72 | 15 | 16.5 | 1.5, 155 | 24 | 2 | 10.6 | 1.2, 88.4 | 19 | 2 | .. | .. | 16 | 1 | .. | .. |
| Term & WHZ <-2 | 667 | 133 | 16.9 | 3.4, 78.7 | 324 | 42 | 15.2 | 3.4, 65.5 | 392 | 60 | 13.6 | 2.8, 63.6 | 314 | 31 | 12.2 | 2.0, 70.3 |
| PTB & WHZ <-2 | 70 | 19 | 31.1 | 3.1, 251 | 14 | 3 | .. | .. | 14 | 3 | .. | .. | 11 | 1 | .. | .. |
| BF & MUAC ≥125 | 969 | 49 | 3.2 | 1.4, 7.4 | 2,428 | 59 | 2.7 | 1.4, 5.0 | 3,374 | 51 | 2.2 | 1.0, 4.9 | 322 | 0 | 1.9 | 0.6, 5.9 |
| No BF & MUAC ≥125 | 50 | 3 | 6.4 | 1.6, 24.4 | 435 | 22 | 5.5 | 2.1, 14.0 | 1,810 | 70 | 4.7 | 2.4, 9.1 | 1,837 | 118 | 4.0 | 2.0, 8.2 |
| BF & MUAC <125 | 2,308 | 361 | 17.0 | 8.6, 33.1 | 1,390 | 126 | 19.2 | 10.6, 34.5 | 1,268 | 74 | 21.7 | 10.0, 46.4 | 60 | 2 | 24.5 | 8.2, 71.0 |
| No BF & MUAC <125 | 264 | 41 | 35.6 | 13.2, 92.2 | 511 | 71 | 40.7 | 20.1, 80.8 | 794 | 105 | 46.6 | 22.9, 92.4 | 131 | 25 | 53.2 | 19.8, 136 |
| NBW & MUAC ≥125 | 886 | 46 | 2.8 | 1.5, 5.0 | 1,875 | 52 | 3.0 | 2.1, 4.3 | 3,309 | 80 | 3.2 | 2.4, 4.4 | 1,463 | 84 | 3.5 | 2.2, 5.6 |
| NBW & MUAC <125 | 45 | 4 | 7.7 | 0.7, 77.2 | 228 | 3 | 5.7 | 1.5, 21.6 | 514 | 4 | 4.3 | 1.2, 15.5 | 82 | 3 | 3.2 | 0.3, 31.1 |
| LBW & MUAC <125 | 2,110 | 323 | 16.4 | 11.7, 22.9 | 966 | 107 | 20.4 | 15.3, 27.1 | 1,110 | 103 | 25.4 | 15.7, 40.8 | 105 | 18 | 31.6 | 15.0, 65.2 |
| LBW & MUAC <125 | 240 | 47 | 20.9 | 7.8, 54.7 | 303 | 23 | 23.9 | 11.8, 47.9 | 304 | 18 | 27.4 | 9.5, 76.3 | 23 | 2 | 31.3 | 5.8, 152 |
| Term & MUAC ≥125 | 706 | 49 | 2.8 | 1.4, 5.5 | 827 | 40 | 3.0 | 2.0, 4.6 | 1,117 | 70 | 3.2 | 2.3, 4.7 | 1,109 | 84 | 3.5 | 2.0, 6.0 |
| PTB & MUAC ≥-125 | 26 | 0 | 0.4 | 0.0, 153 | 24 | 1 | 0.9 | 0.0, 43.2 | 23 | 1 | 2.1 | 0.1, 29.5 | 25 | 2 | 4.9 | 0.1, 160 |
| Term & MUAC <125 | 1,763 | 293 | 15.7 | 10.6, 23.3 | 250 | 49 | 22.2 | 14.5, 33.9 | 247 | 56 | 31.4 | 14.8, 65.6 | 61 | 16 | 44.2 | 14.0, 131 |
| PTB & MUAC <125 | 140 | 39 | 49.1 | 13.5, 162 | 24 | 4 | 38.5 | 6.1, 207 | 10 | 4 | .. | .. | 2 | 0 | .. | .. |

<sup>1</sup> Risk = predicted absolute risk of dying within 1 month per 1,000 child months modelled using generalized linear mixed effects models accounting for repeated measures in each child, clustering within studies, and time at risk
Abbreviations: BF = breastfeeding; HAZ = height-for-age Z-score; LBW= low birthweight (<2500g); MUAC = mid-upper arm circumference; NBW = normal birthweight (≥2500g); PTB = preterm birth (<37 weeks of gestation at birth); WAZ = weight-for-age Z-score; WHZ= weight-for-height Z-score

## DISCUSSION

Among 75,287 children under five years old in 33 pooled cohorts across a range of low resource settings, four readily assessable child-level characteristics – age less than 24 months, low weight-for-age, history of low birthweight or preterm birth, and not breastfeeding in the immediate period preceding – identified children at high risk of mortality. The highest mortality risk (GP studies) was seen at the youngest ages, with a relatively low risk of death beyond two years of age. Young children with even a moderate anthropometric deficit and an additional risk exposure (LBW, PTB or not breastfeeding) were at very high risk.

Our analyses are consistent with other reports that most child deaths occur in the first days of life^2,5^ and that mortality declines rapidly during the first year. Despite these data, infants under twelve months of age are not consistently prioritized in health and nutrition programmes^14^, though some efforts are trying to address this.^15,16^ In contrast to WAZ, WHZ identified mortality risk better with increasing age, but showed poor discrimination among infants as reported elsewhere.^17,18^ An important finding was that concurrent stunting and underweight among infants was associated with increased mortality; whereas, after 12 months of age, concurrent stunting was associated with a lower mortality risk. This possibly reflects specific vulnerabilities associated with small size at birth that are largely lost by one year and points to the need for sustained interventions throughout the first 12 months of life for LBW infants. The mortality risk associated with low MUAC was relatively stable between 1-24 months of age reflecting the balance between older children meeting MUAC criteria being relatively more wasted while their mortality risks tending to decline with increasing age. In our analyses, using cut-offs of <110mm and <120mm for severe and moderate wasting in infants <6 months of age (mentioned in WHO 2023 Wasting guidelines) did not substantially change mortality risks compared to 115mm and 125mm. Nevertheless, it might have implications for caseload and choice of intervention.^19^ Further analyses of the pooled dataset will examine the performance of anthropometric criteria.

As noted by others, children born preterm or with LBW had a higher mortality risk^20–23^. In our analyses, mortality was high throughout the first year of life and remained increased until 24 months of age. The addition of information about LBW, PTB, or not breastfeeding to anthropometry added considerably to the identification of infants at high and lower mortality risk. Between birth and 24 months, children in the GP population who were even moderately underweight but had a history of LBW had increased mortality risk. Notably however, children who were born LBW but who were not underweight, i.e., who had caught up, were not at increased risk. Infants who were moderately underweight and not breastfed were similarly at increased mortality risk. In these infants, recovering or maintaining WAZ probably reflects general nurturing care and health, in addition to appropriate breastfeeding. In A-S and I-S studies, only the combination of any low anthropometric status and not being breastfed carried an additional risk. These analyses demonstrate how the context and inclusion criteria of individual studies needs to be considered when interpreting the performance of risk indicators.

Reported symptoms of diarrhea and LRTI did not add to the predicted mortality risk in the GP and A-S populations. These conditions clearly contribute to child mortality, but the value of self-reported illness is limited. Diarrhea episodes were likely mild and thus did not pose a measurable mortality risk. Pneumonia is similarly difficult to diagnose, and commonly used syndromic definitions have low specificity. There were also few deaths in these children leading to imprecision and probably an underestimation of the risks associated with pneumonia^24^. In I-S studies, the hospitalization period accounted for approximately 50% deaths occurring during 6-months of observation; about 50% post-discharge deaths also occurred within one month of discharge, highlighting both the risks of serious conditions requiring admission and the continued mortality risk in children that persists after discharge from acute care. ^25,26^

Strengths of the analyses include a large sample size originating from 33 relatively recent studies conducted in different regions. The analytical techniques were robust and, different from previous analyses^27,28^, accounted for repeated measures, clustering within studies as well as differences between measurement intervals. Importantly, we report predicted absolute risks by age groups within the first five years of age. Several analyses have previously reported relative risk measures, such as a hazard ratio, and have not generally considered the effects of age^29,30^. While a relative risk informs about the magnitude of association between an exposure and an outcome, it neglects the magnitude of the problem, i.e. the absolute risk of dying in different populations and age groups.

Limitations of the analyses include that studies in the pooled dataset were not necessarily representative of countries or specific populations. Eligibility criteria of original studies may also have skewed the age and exposure distribution which should be considered when interpreting the findings. A paucity of data in the first week of life limited our ability to examine early life risk exposures in more detail.

The analyses demonstrate that assessable, child-level characteristics can be used to identify young children at high and lower risk of mortality. Individual child-centred risk assessment could improve care and outcomes of high-risk children while improving management and efficiencies for lower-risk children. To mitigate risks, low birth weight and preterm infants can be identified at birth and immediately followed-up and supported over a 12-month period to optimise care and nutrition. Low weight-for-age routinely identified at facility or community health visits should prompt the identification of other concurrent risks such as LBW, PTB or not breastfeeding. This would identify children needing closer follow-up and ensure, at minimum, continued monitoring and that vaccinations are received, and mothers are well and able to breastfeed. Young children being discharged from hospital could be actively followed-up in communities and facilities through the first month post-discharge. We need to learn how treatment of specific conditions such as pneumonia or diarrhoea may differ by risk status.

Finally, it is important to recognise that treatment or support for a high-risk child may not primarily target the exposure that identifies the child as being vulnerable. For example, it is not possible to change the birth weight or gestational age. However, it is possible to implement interventions to modify their risk such as improved support and follow-up care. A strength of the findings is their relative simplicity that point to an opportunity and paradigm shift toward more child-centred and targeted care. Avoiding the temptation to over-complicate risk assessments will be essential for programmes.

More effective targeting of interventions to those children at highest risk, while de-escalating care for those at significantly lower risk, offers enormous opportunity. In settings where resources are constrained, it is imperative that we optimize the efficiency of programmes while reducing mortality and morbidity as effectively as possible. These data suggest that simple risk assessment tools that are predicated on individual child characteristics, in addition to the severity of illnesses, may allow for differentiation in care and approaches that further improve child survival, health and development.

## Authors’ contributions

Conceptualization: Catherine Schwinger, Nigel Rollins, Rajiv Bahl, Tor Strand; Design: all WHO-RSWG members; Statistical analysis: Catherine Schwinger, Tor Strand, Ayushi, James A. Berkley; Data acquisition and interpretation: all WHO-RSWG members; Drafting the manuscript: Catherine Schwinger, Nigel Rollins, James A. Berkley, Tor Strand, Judd L. Walson; Reviewing the manuscript: all WHO-RSWG members.

## Data Availability

Principle Investigators of the individual studies can be contacted for a request to access their data.

## Acknowledgments

We thank all study participants and their families for their valuable time in the studies; the teams of each individual study for conducting the study; Tejshri Shah for initial coordination of the working group; and Håkon Gjessing for helpful discussions.

