## Supplementary Material for "Infant- and child-level predictors of mortality in low-resource settings: the WHO Child Mortality Risk Stratification Multi-Country Pooled Cohort"

### Table S1: Study characteristics

| Nr | Short title | Main publication (s) PMID | Country(ies) of implementation | Years of implementation | Study design | Birth cohort[1] | Scheduled study visits | Sample size |
|---|---|---|---|---|---|---|---|---|
| Studies with enrolment not based on anthropometry or the presence of illness (GP)[2] | | | | | | | | |
| 1 | AMANHI Pakistan | 29163938, 34999881 | Pakistan | 2014-2016 | Observational | Yes | <72h, 1-6 days and 42-60 days post-partum | 1,138 |
| 2 | AMANHI Bangladesh | 29163938, 34999881 | Bangladesh | 2014-2016 | Experimental | Yes | <72h, 1-6 days and 42-60 days post-partum | 938 |
| 3 | CHILD 2 | 32518499, 30718805, 29472265 | Tanzania | 2007-2009 | Experimental | Yes | Monthly for 18 months | 2,398 |
| 4 | PNS | 17409323, 4151973 | Tanzania | 2001-2004 | Experimental | Yes | Monthly for 18 months | 7,956 |
| 5 | CRI study | 29099848, 3693507, 17916587 | India | 2002-2006 | Observational | Yes | Bi-weekly up to 26 months | 373 |
| 6 | MAL-ED India | 25305300 | India | 2010-2012 | Observational | Yes | Bi-weekly up to 24 months | 247 |
| 7 | Low BW | 18297930, 15490710 | Burkina Faso | 2004-2005 | Observational | Yes | Monthly for 12 months | 1,103 |
| 8 | ILiNS-ZINC | 25816354, 25521188, 6362661 | Burkina Faso | 2010-2012 | Experimental | No | Weekly for 9 months, and at 12,15 and 18 months of age | 3,220 |
| 9 | ZINC 720 | 27489011 | Burkina Faso | 2010-2012 | Experimental | No | Weekly up to 48 weeks | 6,245 |
| 10 | LAO zinc | 32153900, 30580974, 32612816 | Lao | 2015-2017 | Experimental | No | Weekly up to 9 months (32-40 weeks) | 3,406 |
| 11 | MAL-ED Bangladesh | 25305298 | Bangladesh | 2010-2012 | Observational | Yes | Bi-weekly up to 24 months | 265 |
| 12 | Zambia Study | 29689045, 28588962 | Zambia | 2014-2015 | Experimental | No | After 1 and 2 years | 526 |
| 13 | South Africa Study | 37058529 | South Africa | 2018-2021 | Experimental | No | Monthly up to 2 years of age | 386 |
| 14 | PROMIS Mali | 5343313, 6711497 | Mali | 2015 | Experimental | No | Monthly up to 18 months | 1,132 |
| 15 | PROMIS Burkina Faso | 31454347 | Burkina Faso | 2015 | Experimental | No | Monthly up to 18 months | 2,113 |
| 16 | PM2A Guatemala | 30184223 | Guatemala | 2011-2012 | Experimental | No | At age 1, 4, 6, 9, 12, 18, 24 months | 4,212 |
| 17 | ROSE | 34375336 | Niger | 2014-2015 | Experimental | No | Monthly up to age 24 months | 2,507 |
| 18 | MISAME I | 18996870 | Burkina Faso | 2004-2006 | Experimental | Yes | Monthly for 12 months | 1,221 |
| 19 | MISAME II | 19812173 | Burkina Faso | 2006-2008 | Experimental | Yes | Monthly for 12 months | 1,084 |

| | | | | | | | | |
|---|---|---|---|---|---|---|---|---|
| 20 | MISAME III | 36745684 | Burkina Faso | 2019-2020 | Experimental | Yes | Monthly for 12 months | 1,608 |
| 21 | Keneba | 26559544, 30753251 | Gambia | 2005-2016 | Observational | Yes | Monthly (on average) up to age 5 years | 560 |
| 22 | Pemba | 29163938, 34999881 | Tanzania | 2014-2018 | Observational | Yes | <72h, 1-6 days and 42-60 days post-partum | 4,109 |
| 23 | CARING | 25886587, 28911749 | India | 2013-2015 | Experimental | Yes | <72 h and at 3, 6, 9, 12, and 18 months after birth | 2,993 |
| Studies with enrolment based on anthropometric deficit (A-S)[2] | | | | | | | | |
| 24 | 10% milk RUTF | 20980648 | Malawi | 2008-2009 | Experimental | No | Up to 4 bi-weekly visits | 1,873 |
| 25 | Abx for SAM | 23363496 | Malawi | 2009-2011 | Experimental | No | Up to 6 bi-weekly visits | 2,742 |
| 26 | 3 foods for MAM | 22170366, 23256140, 25419681 | Malawi | 2009-2011 | Experimental | No | Up to 6 bi-weekly visits | 2,707 |
| 27 | COMPAS | 29690916, 32645109 | Kenya, South Sudan | 2017-2018 | Experimental | No | Weekly or bi-weekly | 4,072 |
| 28 | HI MAM | 33963734 | Sierra Leone | 2018-2020 | Experimental | No | After 6, 12 and 24 weeks | 1,286 |
| 29 | DIVIDS | 21628364 | India | 2007-2010 | Experimental | Yes | Monthly up to 6 months | 2,079 |
| Studies with enrolment based on the presence of illness (I-S)[2] | | | | | | | | |
| 30 | CHAIN | 35427524 | Kenya, Uganda, Malawi, Burkina Faso, Bangladesh, Pakistan | 2016-2019 | Observational | No | 45 and 90 days after discharge | 3,101 |
| 31 | SMART 0-6 Phase1 | 37182535 | Uganda | 2018 | Observational | No | Every 2 months for 6 months | 2,647 |
| 32 | SMART 6-60 Phase0 | 26608641 | Uganda | 2017 | Observational | No | Every 2 months for 6 months | 1,273 |
| 33 | SMART 6-60 Phase1 | 37182535 | Uganda | 2018 | Observational | No | Every 2 months for 6 months | 3,767 |

[1]As birth cohorts, we included studies enrolling women during pregnancy or neonates up until 17 days after births

[2] A study was categorized as General Population (GP) when enrolment to the original study was not based on an anthropometric deficit or the presence of an illness even if some additional inclusion criteria may have been implemented, and as Anthropometry-Selected (A-S) or Illness-Selected (I-S) if the original study enrolled children based on an anthropometric deficit (low WHZ, MUAC or low birthweight) or admitted to hospital due to the presence of signs or symptoms of an acute illness, respectively.

Abbreviations: BW = birthweight; MAM = moderate acute malnutrition; RUTF = ready to use therapeutic food; SAM = severe acute malnutrition·

**Table S2 – Absolute risk of death per 1,000 child months by age group and by individual exposures in General Population (GP) cohorts, i·e·, enrolment not on the basis of anthropometric deficit or the presence of illness for all children with death age available**

| | <6 months | | | | 6-11 months | | | | 12-23 months | | | | 24-59 months | | | |
|---|---|---|---|---|---|---|---|---|---|---|---|---|---|---|---|---|
| **Indicator** | **N** | **Death** | **Risk[1]** | **95% CI** | **N** | **Death** | **Risk[1]** | **95% CI** | **N** | **Death** | **Risk[1]** | **95% CI** | **N** | **Death** | **Risk[1]** | **95% CI** |
| **ALL** | 142,010 | 637 | 2·0 | 1·3, 3·2 | 135,403 | 278 | 1·0 | 0·6, 1·5 | 149,241 | 166 | 0·7 | 0·5, 1·1 | 35,239 | 12 | 0·5 | 0·2, 0·9 |
| **WAZ** | | | | | | | | | | | | | | | | |
| ≥-2 | 113,637 | 285 | 1·0 | 0·6, 1·7 | 100,340 | 140 | 0·6 | 0·3, 1·0 | 95,032 | 61 | 0·3 | 0·2, 0·6 | 22,097 | 3 | 0·2 | 0·1, 0·3 |
| <-2, ≥-3 | 10,015 | 56 | 2·4 | 1·4, 4·3 | 16,776 | 53 | 1·5 | 0·9, 2·5 | 22,867 | 33 | 0·9 | 0·5, 1·6 | 6,380 | 3 | 0·5 | 0·3, 1·1 |
| <-3 | 4,535 | 134 | 12·8 | 7·5, 21·9 | 6,612 | 54 | 6·0 | 3·5, 10·1 | 8,113 | 36 | 2·8 | 1·5, 5·0 | 1,929 | 4 | 1·3 | 0·6, 2·6 |
| Missing | 13,823 | 162 | 5·3 | 3·1, 9·1 | 11,675 | 31 | 4·2 | 2·5, 7·1 | 23,229 | 36 | 3·3 | 1·8, 6·0 | 4,833 | 2 | 2·6 | 1·3, 5·4 |
| **HAZ** | | | | | | | | | | | | | | | | |
| ≥-2 | 106,952 | 274 | 1·1 | 0·6, 1·8 | 95,668 | 146 | 0·6 | 0·4, 1·1 | 74,782 | 61 | 0·4 | 0·2, 0·7 | 16,317 | 1 | 0·2 | 0·1, 0·4 |
| <-2, ≥-3 | 13,608 | 77 | 2·5 | 1·4, 4·3 | 20,357 | 56 | 1·2 | 0·7, 2·0 | 34,029 | 31 | 0·6 | 0·3, 1·0 | 9,271 | 3 | 0·3 | 0·1, 0·5 |
| <-3 | 6,125 | 117 | 8·3 | 4·8, 14·4 | 7,428 | 45 | 3·4 | 2·0, 5·8 | 16,788 | 35 | 1·4 | 0·8, 2·5 | 4,672 | 6 | 0·6 | 0·3, 1·1 |
| Missing | 15,314 | 169 | 5·4 | 3·1, 9·3 | 11,944 | 31 | 4·2 | 2·4, 7·2 | 23,618 | 38 | 3·2 | 1·7, 5·9 | 4,977 | 2 | 2·5 | 1·2, 5·1 |
| **WHZ** | | | | | | | | | | | | | | | | |
| ≥-2 | 117,078 | 333 | 1·3 | 0·8, 2·1 | 110,901 | 183 | 0·7 | 0·4, 1·2 | 113,137 | 89 | 0·4 | 0·3, 0·7 | 28,149 | 8 | 0·3 | 0·1, 0·4 |
| <-2, ≥-3 | 5,494 | 28 | 2·2 | 1·2, 4·0 | 9,428 | 38 | 1·5 | 0·9, 2·6 | 9,784 | 17 | 1·1 | 0·6, 2·0 | 1,784 | 0 | 0·7 | 0·3, 1·7 |
| <-3 | 2,402 | 29 | 3·9 | 2·1, 7·2 | 3,000 | 26 | 4·8 | 2·8, 8·3 | 2,626 | 21 | 6·1 | 3·3, 11·3 | 305 | 2 | 7·6 | 3·4, 17·1 |
| Missing | 17,025 | 247 | 7·5 | 4·5, 12·5 | 12,068 | 31 | 4·9 | 2·9, 8·1 | 23,670 | 38 | 3·1 | 1·8, 5·6 | 4,999 | 2 | 2·0 | 1·0, 4·1 |
| **MUAC** | | | | | | | | | | | | | | | | |
| ≥125 | 53,279 | 116 | 1·3 | 0·8, 2·2 | 88,415 | 148 | 1·0 | 0·6, 1·6 | 99,126 | 85 | 0·7 | 0·4, 1·2 | 15,817 | 6 | 0·5 | 0·3, 0·9 |
| <125, ≥115 | 14,732 | 62 | 2·1 | 1·2, 3·5 | 8,863 | 40 | 2·3 | 1·4, 3·8 | 6,660 | 18 | 2·5 | 1·4, 4·6 | 340 | 1 | 2·8 | 1·3, 6·0 |
| <115 | 17,965 | 228 | 5·2 | 3·2, 8·4 | 1,870 | 13 | 7·0 | 4·2, 117 | 993 | 16 | 9·4 | 4·8, 18·0 | 65 | 2 | 12·5 | 5·3, 29·4 |
| Missing | 40,354 | 195 | 3·7 | 2·3, 6·1 | 21,734 | 48 | 3·6 | 2·2, 5·9 | 23,179 | 37 | 3·6 | 2·1, 6·2 | 4,519 | 2 | 3·5 | 1·8, 6·8 |
| **MUAC (0-6 mo)** | | | | | | | | | | | | | | | | |
| ≥120 | 63,022 | 156 | 1·4 | .. | .. | .. | .. | .. | .. | .. | .. | .. | .. | .. | .. | .. |
| <120, ≥110 | 10,175 | 51 | 1·9 | .. | .. | .. | .. | .. | .. | .. | .. | .. | .. | .. | .. | .. |
| <110 | 12,779 | 199 | 5·9 | .. | .. | .. | .. | .. | .. | .. | .. | .. | .. | .. | .. | .. |
| Missing | 40,354 | 195 | 5·3 | .. | .. | .. | .. | .. | .. | .. | .. | .. | .. | .. | .. | .. |
| **any BF** | | | | | | | | | | | | | | | | |
| No | 993 | 14 | 3·7 | 1·5, 9·1 | 3,191 | 15 | 2·0 | 0·9, 4·3 | 23,239 | 20 | 1·1 | 0·5, 2·4 | 17,491 | 5 | 0·6 | 0·2 ,1·5 |
| Yes | 91,028 | 132 | 0·6 | 0·3, 1·3 | 94,053 | 156 | 0·5 | 0·3, 1·1 | 82,120 | 72 | 0·4 | 0·2, 0·9 | 4,191 | 4 | 0·4 | 0·2, 0·8 |
| Missing | 20,044 | 196 | 2·8 | 1·3, 5·8 | 24,921 | 69 | 1·4 | 0·7, 2·9 | 32,381 | 54 | 0·7 | 0·3, 1·5 | 3,499 | 1 | 0·3 | 0·1, 0·8 |
| **LBW** | | | | | | | | | | | | | | | | |
| No | 90,711 | 345 | 1·3 | 0·7, 2·5 | 70,255 | 129 | 0·7 | 0·4, 1·4 | 44,349 | 37 | 0·4 | 0·2, 0·8 | 19,383 | 2 | 0·2 | 0·1, 0·5 |
| Yes | 11,625 | 213 | 6·9 | 3·5, 13·3 | 8,252 | 35 | 2·0 | 1·0, 4·0 | 5,283 | 10 | 0·6 | 0·2, 1·3 | 2,542 | 0 | 0·2 | 0·1, 0·5 |
| Missing | 15,571 | 32 | 2·0 | 0·9, 4·2 | 16,978 | 19 | 1·6 | 0·8, 3·4 | 29,558 | 19 | 1·3 | 0·6, 3·0 | 2,854 | 2 | 1·1 | 0·4, 2·9 |
| **PTB** | | | | | | | | | | | | | | | | |

| | | | | | | | | | | | | | | | | |
|---|---|---|---|---|---|---|---|---|---|---|---|---|---|---|---|---|
| No | 87,671 | 398 | 2·4 | 1·5, 3·9 | 66,502 | 138 | 1·2 | 0·8, 2·0 | 39,066 | 42 | 0·6 | 0·4, 1·1 | 10,678 | 2 | 0·3 | 0·2, 0·6 |
| Yes | 13,788 | 170 | 8·2 | 4·9, 13·5 | 10,558 | 36 | 3·0 | 1·8, 5·1 | 5,981 | 12 | 1·1 | 0·6, 2·1 | 937 | 0 | 0·4 | 0·2, 0·9 |
| Missing | 13,704 | 9 | 2·1 | 0·9, 4·8 | 15,469 | 14 | 2·3 | 1·1, 4·9 | 28,971 | 17 | 2·7 | 1·2, 5·9 | 2,364 | 2 | 3·0 | 1·2, 8·3 |
| **Diarrhea** | | | | | | | | | | | | | | | | |
| No | 81,159 | 188 | 1·0 | 0·5, 1·8 | 81,323 | 143 | 0·8 | 0·4, 1·4 | 103,961 | 83 | 0·6 | 0·3, 1·2 | 23,086 | 8 | 0·5 | 0·3, 1·0 |
| Yes | 4,534 | 18 | 2·1 | 1·0, 4·4 | 9,062 | 32 | 1·5 | 0·8, 2·9 | 8,693 | 12 | 1·1 | 0·5, 2·2 | 1,415 | 1 | 0·8 | 0·3, 2·0 |
| Missing | 43,266 | 276 | 3·0 | 1·6, 5·6 | 40,422 | 87 | 1·9 | 1·0, 3·5 | 26,512 | 53 | 1·2 | 0·6· 2·3 | 676 | 1 | 0·8 | 0·4, 1·6 |
| **LRTI** | | | | | | | | | | | | | | | | |
| No | 33,407 | 39 | 0·7 | 0·4, 1·5 | 40,094 | 58 | 0·8 | 0·4, 1·5 | 61,069 | 59 | 0·9 | 0·5, 1·6 | 11,453 | 3 | 0·9 | 0·5, 1·9 |
| Yes | 1,611 | 2 | 1·2 | 0·3, 4·1 | 2,105 | 5 | 1·0 | 0·4, 2·7 | 1,055 | 0 | 0·8 | 0·2, 4·4 | 72 | 0 | 0·7 | 0·1, 9·3 |
| Missing | 35,476 | 170 | 2·1 | 1·1, 3·9 | 41,955 | 84 | 1·3 | 0·7, 2·4 | 29,882 | 57 | 0·8 | 0·4, 1·6 | 1,243 | 4 | 0·5 | 0·3, 1·1 |

[1] Risk = predicted absolute risk of dying within 1 month per 1,000 child months modelled in mixed linear regression models accounting for repeated measures in each child, clustering within studies, and time at risk

Abbreviations: BF = breastfeeding; HAZ = height-for-age Z-score; LBW= low birthweight (<2500g); LRTI = acute lower respiratory infection; MUAC = mid-upper arm circumference; PTB = preterm birth (<37 weeks of gestation at birth); WAZ = weight-for-age Z-score; WHZ= weight-for-height Z-score·

**Table S3 – Absolute risk of death per 1,000 child months by age group and by individual exposures in General Population (GP) cohorts, i·e·, enrolment not on the basis of anthropometric deficit or the presence of illness**

| | <6 months | | | | 6-11 months | | | | 12-23 months | | | | 24-59 months | | | |
|---|---|---|---|---|---|---|---|---|---|---|---|---|---|---|---|---|
| **Indicator** | **N** | **Death** | **Risk[1]** | **95% CI** | **N** | **Death** | **Risk[1]** | **95% CI** | **N** | **Death** | **Risk[1]** | **95% CI** | **N** | **Death** | **Risk[1]** | **95% CI** |
| **ALL** | 142,059 | 686 | 2·0 | 1·3, 3·2 | 135,464 | 333 | 1·1 | 0·7, 1·7 | 150,232 | 205 | 0·9 | 0·5, 1·4 | 35,450 | 14 | 0·6 | 0·3, 1·2 |
| **WAZ** | | | | | | | | | | | | | | | | |
| ≥-2 | 113,654 | 302 | 1·0 | 0·6, 1·7 | 100,372 | 172 | 0·6 | 0·4, 1·1 | 95,054 | 83 | 0·4 | 0·2, 0·7 | 22,097 | 3 | 0·3 | 0·1, 0·5 |
| <-2, ≥-3 | 10,019 | 60 | 2·3 | 1·2, 4·1 | 16,784 | 61 | 1·5 | 0·9, 2·7 | 22,872 | 38 | 1·0 | 0·6, 1·9 | 6,382 | 5 | 0·7 | 0·3, 1·4 |
| <-3 | 4,540 | 139 | 11·0 | 6·2, 19·5 | 6,623 | 65 | 5·7 | 3·2, 10·0 | 8,120 | 43 | 2·9 | 1·6, 5·5 | 1,929 | 4 | 1·5 | 0·7, 3·1 |
| Missing | 13,846 | 185 | 5·7 | 3·2, 10·1 | 11,679 | 35 | 4·7 | 2·7, 8·4 | 23,234 | 41 | 4·0 | 2·1, 7·4 | 4,833 | 2 | 3·3 | 1·6, 6·8 |
| **HAZ** | | | | | | | | | | | | | | | | |
| ≥-2 | 106,968 | 290 | 1·0 | 0·6, 1·8 | 95,706 | 184 | 0·7 | 0·4, 1·3 | 74,805 | 84 | 0·5 | 0·3, 0·9 | 16,319 | 3 | 0·4 | 0·2, 0·7 |
| <-2, ≥-3 | 13,613 | 82 | 2·5 | 1·4, 4·5 | 20,366 | 65 | 1·3 | 0·7, 2·2 | 34,033 | 35 | 0·6 | 0·3, 1·2 | 9,271 | 3 | 0·3 | 0·2, 0·6 |
| <-3 | 6,130 | 122 | 7·4 | 4·1, 13·3 | 7,432 | 49 | 3·4 | 1·9, 6·0 | 16,795 | 42 | 1·6 | 0·8, 2·9 | 4,672 | 6 | 0·7 | 0·4, 1·4 |
| Missing | 15,337 | 192 | 5·7 | 3·2, 10·1 | 11,948 | 35 | 4·6 | 2·6, 8·3 | 23,623 | 43 | 3·8 | 2·0, 7·1 | 4,977 | 2 | 3·1 | 1·5, 6·4 |
| **WHZ** | | | | | | | | | | | | | | | | |
| ≥-2 | 117,096 | 351 | 1·2 | 0·7, 2·1 | 110,936 | 218 | 0·8 | 0·5, 1·4 | 113,161 | 113 | 0·5 | 0·3, 0·9 | 28,149 | 8 | 0·3 | 0·2, 0·6 |
| <-2, ≥-3 | 5,496 | 30 | 2·1 | 1·1, 3·9 | 9,440 | 50 | 1·8 | 1·0, 3·1 | 9,789 | 22 | 1·5 | 0·8, 2·8 | 1,786 | 2 | 1·3 | 0·6, 2·7 |
| <-3 | 2,403 | 30 | 3·5 | 1·8, 6·7 | 3,004 | 50 | 4·8 | 2·7, 8·5 | 2,631 | 26 | 6·5 | 3·4, 12·4 | 305 | 2 | 8·7 | 3·9, 19·6 |
| Missing | 17,053 | 275 | 7·5 | 4·3, 13·0 | 12,072 | 35 | 5·3 | 3·0, 9·2 | 23,675 | 43 | 3·7 | 2·0, 6·8 | 4,999 | 2 | 2·6 | 1·3, 5·3 |
| **MUAC** | | | | | | | | | | | | | | | | |
| ≥125 | 53,289 | 126 | 1·3 | 0·8, 2·2 | 88,451 | 184 | 1·1 | 0·7, 1·8 | 99,155 | 114 | 1·0 | 0·6, 1·6 | 15,819 | 8 | 0·8 | 0·5, 1·5 |
| <125, ≥115 | 14,733 | 63 | 2·1 | 1·2, 3·6 | 8,872 | 49 | 2·4 | 1·4, 4·1 | 6,661 | 19 | 2·8 | 1·6, 7·1 | 340 | 1 | 3·4 | 1·6, 7·1 |
| <115 | 17,978 | 241 | 5·1 | 3·1, 8·5 | 1,876 | 19 | 7·3 | 4·3, 12·4 | 996 | 19 | 10·5 | 5·5, 20·0 | 65 | 2 | 15·0 | 6·5, 33·9 |
| Missing | 40,377 | 218 | 4·2 | 2·5, 7·0 | 21,738 | 52 | 4·2 | 2·5, 7·1 | 23,184 | 42 | 4·3 | 2·4, 7·6 | 4,519 | 2 | 4·4 | 2·2, 8·5 |
| **MUAC (0-6 mo)** | | | | | | | | | | | | | | | | |
| ≥120 | 63,032 | 166 | 1·4 | 0·8, 2·5 | .. | .. | .. | .. | .. | .. | .. | .. | .. | .. | .. | .. |
| <120, ≥110 | 10,177 | 53 | 2·0 | 1·1, 3·7 | .. | .. | .. | .. | .. | .. | .. | .. | .. | .. | .. | .. |
| <110 | 12,791 | 211 | 5·7 | 3·3, 10·0 | .. | .. | .. | .. | .. | .. | .. | .. | .. | .. | .. | .. |
| Missing | 40,377 | 218 | 5·6 | 3·1, 9·9 | .. | .. | .. | .. | .. | .. | .. | .. | .. | .. | .. | .. |
| **any BF** | | | | | | | | | | | | | | | | |
| No | 993 | 14 | 3·8 | 1·6, 9·0 | 3,192 | 16 | 2·2 | 1·0, 4·6 | 23,239 | 20 | 1·2 | 0·6, 2·6 | 17,492 | 6 | 0·7 | 0·3 ,1·7 |
| Yes | 91,031 | 135 | 0·7 | 0·3, 1·4 | 94,078 | 181 | 0·6 | 0·3, 1·2 | 82,135 | 87 | 0·5 | 0·2, 1·0 | 4,198 | 5 | 0·4 | 0·2, 0·9 |
| Missing | 20,046 | 218 | 2·9 | 1·4, 6·0 | 24,931 | 73 | 1·5 | 0·7, 3·0 | 32,426 | 60 | 0·8 | 0·4, 1·6 | 3,523 | 1 | 0·4 | 0·2, 0·9 |
| **LBW** | | | | | | | | | | | | | | | | |
| No | 90,736 | 370 | 1·2 | 0·6, 2·5 | 70,278 | 152 | 0·8 | 0·4, 1·6 | 45,038 | 54 | 0·5 | 0·2, 1·0 | 19,383 | 2 | 0·3 | 0·2, 0·7 |
| Yes | 11,646 | 234 | 6·2 | 3·1, 12·6 | 8,258 | 41 | 2·4 | 1·2, 4·9 | 5,498 | 16 | 0·9 | 0·4, 2·1 | 2,542 | 0 | 0·4 | 0·1, 0·9 |
| Missing | 15,572 | 33 | 1·8 | 0·8, 4·0 | 16,979 | 20 | 1·6 | 0·7, 3·4 | 25,591 | 20 | 1·4 | 0·6, 3·2 | 2,854 | 2 | 1·2 | 0·4, 3·2 |
| **PTB** | | | | | | | | | | | | | | | | |
| No | 87,704 | 431 | 2·5 | 1·4, 4·4 | 66,526 | 162 | 1·5 | 0·8, 2·6 | 39,083 | 59 | 0·9 | 0·5, 1·6 | 10,678 | 2 | 0·5 | 0·3, 1·0 |
| Yes | 13,804 | 186 | 8·6 | 4·9, 15·2 | 10,564 | 42 | 3·8 | 2·1, 6·8 | 5,987 | 18 | 1·6 | 0·8, 3·2 | 937 | 0 | 0·7 | 0·3, 1·6 |
| Missing | 13,704 | 9 | 2·3 | 1·0, 5·6 | 15,471 | 14 | 2·9 | 1·4, 6·2 | 28,973 | 19 | 3·6 | 1·6, 8·2 | 2,364 | 2 | 4·5 | 1·6, 12·5 |
| **Diarrhea** | | | | | | | | | | | | | | | | |
| No | 81,172 | 201 | 0·9 | 0·4, 1·7 | 81,344 | 164 | 0·8 | 0·4, 1·6 | 103,989 | 111 | 0·8 | 0·4, 1·6 | 23,095 | 9 | 0·8 | 0·4, 1·6 |
| Yes | 4,535 | 19 | 1·9 | 0·9, 4·0 | 9,064 | 34 | 1·6 | 0·8, 3·1 | 8,697 | 16 | 1·4 | 0·6, 2·9 | 1,416 | 1 | 1·2 | 0·5, 3·0 |
| Missing | 43,301 | 311 | 3·0 | 1·6, 5·8 | 40,454 | 119 | 2·1 | 1·1, 4·0 | 26,519 | 60 | 1·5 | 0·7· 2·9 | 678 | 2 | 1·0 | 0·5, 2·1 |

| | | | | | | | | | | | | | | | | |
|---|---|---|---|---|---|---|---|---|---|---|---|---|---|---|---|---|
| **LRTI** | | | | | | | | | | | | | | | | |
| No | 33,407 | 39 | 0·8 | 0·4, 1·5 | 40,094 | 58 | 0·8 | 0·4, 1·6 | 61,076 | 67 | 0·9 | 0·5, 1·8 | 11,453 | 3 | 1·0 | 0·5, 2·1 |
| Yes | 1,611 | 2 | 1·1 | 0·3, 4·0 | 2,105 | 5 | 1·3 | 0·5, 3·3 | 1,056 | 1 | 1·4 | 0·3, 5·9 | 72 | 0 | 1·6 | 0·2, 14·6 |
| Missing | 35,476 | 193 | 2·4 | 1·3, 4·7 | 41,985 | 114 | 1·6 | 0·8, 3·0 | 29,893 | 68 | 1·0 | 0·5, 2·0 | 1,245 | 6 | 0·7 | 0·3, 1·4 |

[1] Risk = predicted absolute risk of dying within 1 month per 1,000 child months modelled in mixed linear regression models accounting for repeated measures in each child, clustering within studies, and time at risk

Abbreviations: BF = breastfeeding; HAZ = height-for-age Z-score; LBW= low birthweight (<2500g); LRTI = acute lower respiratory infection; MUAC = mid-upper arm circumference; PTB = preterm birth (<37 weeks of gestation at birth); WAZ = weight-for-age Z-score; WHZ= weight-for-height Z-score

**Table S4 – Mortality odds ratio by age group and by individual exposures in General Population (GP) cohorts, i·e·, enrolment not on the basis of anthropometric deficit or the presence of illness**

| | <6 months | | | | 6-11 months | | | | 12-23 months | | | | 24-59 months | | | |
|---|---|---|---|---|---|---|---|---|---|---|---|---|---|---|---|---|
| **Indicator** | N | **Death** | **OR** | **95% CI** | N | **Death** | **OR** | **95% CI** | N | **Death** | **OR** | **95% CI** | N | **Death** | **OR** | **95% CI** |
| **WAZ** | | | | | | | | | | | | | | | | |
| ≥-2 | 113,654 | 302 | Ref | | 100,372 | 172 | Ref | | 95,054 | 83 | Ref | | 22,097 | 3 | Ref | |
| <-2, ≥-3 | 10,019 | 60 | 2·65 | 1·74, 4·04 | 16,784 | 61 | 3·01 | 0·61, 14·9 | 22,872 | 38 | 4·09 | 0·43, 38·6 | 6,382 | 5 | ·· | ·· |
| <-3 | 4,540 | 139 | 19·2 | 13·7, 26·7 | 6,623 | 65 | 4·67 | 0·92, 23·6 | 8,120 | 43 | 6·98 | 0·80, 61·2 | 1,929 | 4 | ·· | ·· |
| Missing | 13,846 | 185 | 5·50 | 3·96, 7·66 | 11,679 | 35 | 8·70 | 1·22, 62·1 | 23,234 | 41 | 9·24 | 0·94, 90·8 | 4,833 | 2 | ·· | ·· |
| **HAZ** | | | | | | | | | | | | | | | | |
| ≥-2 | 106,968 | 290 | Ref | | 95,706 | 184 | Ref | | 74,805 | 84 | Ref | | 16,319 | 3 | Ref | |
| <-2, ≥-3 | 13,613 | 82 | 2·07 | 1·38, 3·09 | 20,366 | 65 | 2·02 | 0·43, 9·45 | 34,033 | 35 | 10·27 | 0·99-106 | 9,271 | 3 | ·· | ·· |
| <-3 | 6,130 | 122 | 9·22 | 6·68, 12·7 | 7,432 | 49 | 4·26 | 0·82, 22·0 | 16,795 | 42 | 10·52 | 1·17, 94·4 | 4,672 | 6 | ·· | ·· |
| Missing | 15,337 | 192 | 5·71 | 4·13, 7·90 | 11,948 | 35 | 6·44 | 0·91, 45·6 | 23,623 | 43 | 9·21 | 0·98, 86·6 | 4,977 | 2 | ·· | ·· |
| **WHZ** | | | | | | | | | | | | | | | | |
| ≥-2 | 117,096 | 351 | Ref | | 110,936 | 218 | Ref | | 113,161 | 113 | Ref | | 28,149 | 8 | Ref | |
| <-2, ≥-3 | 5,496 | 30 | 1·99 | 1·15, 3·43 | 9,440 | 50 | 1·05 | 0·15, 7·15 | 9,789 | 22 | 3·54 | 0·21, 60·4 | 1,786 | 2 | ·· | ·· |
| <-3 | 2,403 | 30 | 1·89 | 0·98, 3·63 | 3,004 | 50 | 13·24 | 1·74, 101 | 2,631 | 26 | 2·09 | 0·17, 26·1 | 305 | 2 | ·· | ·· |
| Missing | 17,053 | 275 | 6·69 | 5·17, 8·65 | 12,072 | 35 | 6·55 | 0·96, 44·4 | 23,675 | 43 | 5·56 | 0·67, 45·9 | 4,999 | 2 | ·· | ·· |
| **MUAC** | | | | | | | | | | | | | | | | |
| ≥125 | 53,289 | 126 | Ref | | 88,451 | 184 | Ref | | 99,155 | 114 | Ref | | 15,819 | 8 | Ref | |
| <125, ≥115 | 14,733 | 63 | 1·09 | 0·55, 2·13 | 8,872 | 49 | 0·94 | 0·15, 6·07 | 6,661 | 19 | 9·78 | 0·60, 158 | 340 | 1 | ·· | ·· |
| <115 | 17,978 | 241 | 3·34 | 19·5, 5·73 | 1,876 | 19 | 4·81 | 0·28, 83·3 | 996 | 19 | 6·57 | 0·43, 100 | 65 | 2 | ·· | ·· |
| Missing | 40,377 | 218 | 7·61 | 4·30, 13·5 | 21,738 | 52 | 2·58 | 0·45, 14·9 | 23,184 | 42 | 8·20 | 0·96, 70·1 | 4,519 | 2 | ·· | ·· |
| **MUAC (0-6 mo)** | | | | | | | | | | | | | | | | |
| ≥120 | 63,032 | 166 | Ref | | | | | | | | | | | | | |
| <120, ≥110 | 10,177 | 53 | 1·28 | 0·74, 2·19 | ·· | ·· | ·· | ·· | ·· | ·· | ·· | ·· | ·· | ·· | ·· | ·· |
| <110 | 12,791 | 211 | 3·92 | 2·57, 5·98 | ·· | ·· | ·· | ·· | ·· | ·· | ·· | ·· | ·· | ·· | ·· | ·· |
| Missing | 40,377 | 218 | 7·29 | 4·59, 11·6 | ·· | ·· | ·· | ·· | ·· | ·· | ·· | ·· | ·· | ·· | ·· | ·· |
| **any BF** | | | | | | | | | | | | | | | | |
| No | 993 | 14 | Ref | | 3,192 | 16 | Ref | | 23,239 | 20 | Ref | | 17,492 | 6 | Ref | |
| Yes | 91,031 | 135 | 0·09 | 0·02, 0·34 | 94,078 | 181 | 0·30 | 0·02 4·57 | 82,135 | 87 | 0·0 | 0·0, 0·04 | 4,198 | 5 | ·· | ·· |
| Missing | 20,046 | 218 | 0·74 | 0·20, 2·70 | 24,931 | 73 | 1·05 | 0·06, 19·4 | 32,426 | 60 | 0·8 | 0·0, 2·38 | 3,523 | 1 | ·· | ·· |
| **LBW** | | | | | | | | | | | | | | | | |
| No | 90,736 | 370 | Ref | | 70,278 | 152 | Ref | | 45,038 | 54 | Ref | | 19,383 | 2 | Ref | |
| Yes | 11,646 | 234 | 6·34 | 4·98, 8·07 | 8,258 | 41 | 1·15 | 0·19, 6·83 | 5,498 | 16 | 9·98 | 0·03, 2936 | 2,542 | 0 | ·· | ·· |
| Missing | 15,572 | 33 | 1·62 | 0·85, 3·11 | 16,979 | 20 | 2·25 | 0·21, 24·0 | 25,591 | 20 | 0·03 | 0·0, 1·09 | 2,854 | 2 | ·· | ·· |
| **PTB** | | | | | | | | | | | | | | | | |
| No | 87,704 | 431 | Ref | | 66,526 | 162 | Ref | | 39,083 | 59 | Ref | | 10,678 | 2 | Ref | |
| Yes | 13,804 | 186 | 4·83 | 3·77, 6·18 | 10,564 | 42 | 4·05 | 0·70, 23·4 | 5,987 | 18 | 12·2 | 0·12, 1285 | 937 | 0 | ·· | ·· |
| Missing | 13,704 | 9 | 0·05 | 0·0, 0·67 | 15,471 | 14 | 5·66 | 0·31, 104 | 28,973 | 19 | 0·44 | 0·02, 12·8 | 2,364 | 2 | ·· | ·· |
| **Diarrhea** | | | | | | | | | | | | | | | | |
| No | 81,172 | 201 | Ref | | 81,344 | 164 | Ref | | 103,989 | 111 | Ref | | 23,095 | 9 | Ref | |
| Yes | 4,535 | 19 | 1·77 | 0·50, 6·25 | 9,064 | 34 | 1·52 | 0·23, 10·2 | 8,697 | 16 | 0·32 | 0·01, 8·66 | 1,416 | 1 | ·· | ·· |
| Missing | 43,301 | 311 | 4·58 | 3·15, 6·66 | 40,454 | 119 | 1·64 | 0·35, 7·67 | 26,519 | 60 | 270 | 23·0, 3167 | 678 | 2 | ·· | ·· |
| **LRTI** | | | | | | | | | | | | | | | | |

| | | | | | | | | | | | | | | | | |
|---|---|---|---|---|---|---|---|---|---|---|---|---|---|---|---|---|
| No | 33,407 | 39 | Ref | | 40,094 | 58 | Ref | | 61,076 | 67 | Ref | | 11,453 | 3 | Ref | |
| Yes | 1,611 | 2 | 5·90 | 0·24, 143 | 2,105 | 5 | 0·07 | 0·00, 13·5 | 1,056 | 1 | - | | 72 | 0 | ·· | ·· |
| Missing | 35,476 | 193 | 7·95 | 3·62, 17·5 | 41,985 | 114 | 0·86 | 0·14, 5·40 | 29,893 | 68 | - | | 1,245 | 6 | ·· | ·· |

··

Abbreviations: BF = breastfeeding; HAZ = height-for-age Z-score; LBW= low birthweight (<2500g); LRTI = acute lower respiratory infection; MUAC = mid-upper arm circumference; OR = Odds ratio; PTB = preterm birth (<37 weeks of gestation at birth); WAZ = weight-for-age Z-score; WHZ= weight-for-height Z-score

**Table S5 – Absolute risk of death per 1,000 child months of weight-for-age Z-score (WAZ) categories by age group and by a) stunting status or b) sex, in General Population (GP) cohorts, i·e·, enrolment not on the basis of anthropometric deficit or the presence of illness**

| | | <6 months | | | | 6-11 months | | | | 12-23 months | | | | 24-59 months | | | |
|---|---|---|---|---|---|---|---|---|---|---|---|---|---|---|---|---|---|
| | **Indicator** | **N** | **Deaths** | **Risk[1]** | **95% CI** | **N** | **Deaths** | **Risk[1]** | **95% CI** | **N** | **Deaths** | **Risk[1]** | **95% CI** | **N** | **Deaths** | **Risk[1]** | **95% CI** |
| **Not stunted** | **WAZ** | | | | | | | | | | | | | | | | |
| | ≥-2 | 101,283 | 257 | 1·1 | 0·6, 1·9 | 86,538 | 145 | 0·7 | 0·4, 1·2 | 68,767 | 60 | 0·4 | 0·2, 0·7 | 15,073 | 1 | 0·3 | 0·1, 0·5 |
| | <-2, ≥-3 | 4,839 | 24 | 1·6 | 0·8, 3·1 | 8·012 | 31 | 1·6 | 0·9, 2·8 | 5,351 | 16 | 1·5 | 0·8, 3·0 | 1,155 | 2 | 1·5 | 0·6, 3·5 |
| | <-3 | 760 | 9 | 3·2 | 1·4, 7·5 | 1,079 | 8 | 4·8 | 2·5, 9·3 | 650 | 8 | 7·2 | 3·0, 17·2 | 82 | 0 | 10·6 | 2·9, 38·8 |
| **Stunted** | **WAZ** | | | | | | | | | | | | | | | | |
| | ≥-2 | 10,908 | 40 | 1·8 | 1·0, 3·0 | 13,553 | 27 | 0·9 | 0·6, 1·5 | 25,900 | 23 | 0·5 | 0·3, 0·8 | 6,885 | 2 | 0·2 | 0·1, 0·5 |
| | <-2, ≥-3 | 5,093 | 36 | 3·7 | 2·2, 6·3 | 8,726 | 30 | 1·9 | 1·2, 3·0 | 17,464 | 21 | 1·0 | 0·5, 1·7 | 5,211 | 3 | 0·5 | 0·2, 1·0 |
| | <-3 | 3,726 | 128 | 17·1 | 10·7, 27·4 | 5,510 | 57 | 7·5 | 4·7, 11·8 | 7,449 | 33 | 3·2 | 1·9, 5·5 | 1,843 | 4 | 1·4 | 0·7, 2·7 |
| **Male** | **WAZ** | | | | | | | | | | | | | | | | |
| | ≥-2 | 56,462 | 152 | 1·1 | 0·7, 1·9 | 48,782 | 89 | 0·8 | 0·5, 1·2 | 45,670 | 44 | 0·5 | 0·3, 0·8 | 10,944 | 1 | 0·3 | 0·2, 0·6 |
| | <-2, ≥-3 | 5,596 | 37 | 2·8 | 1·6, 4·9 | 9,813 | 32 | 1·8 | 1·1, 3·0 | 12,801 | 20 | 1·2 | 0·6, 2·1 | 3,236 | 5 | 0·7 | 0·4, 1·6 |
| | <-3 | 2,818 | 80 | 12·3 | 7·2, 20·7 | 4,317 | 36 | 6·4 | 3·9, 10·6 | 5,098 | 30 | 3·4 | 1·9, 6·1 | 1,079 | 1 | 1·8 | 0·8, 3·7 |
| **Female** | **WAZ** | | | | | | | | | | | | | | | | |
| | ≥-2 | 57,192 | 150 | 1·3 | 0·8, 2·1 | 51,590 | 83 | 0·7 | 0·5, 1·2 | 49,384 | 39 | 0·4 | 0·2, 0·7 | 11,153 | 2 | 0·2 | 0·1, 0·5 |
| | <-2, ≥-3 | 4,423 | 23 | 2·6 | 1·4, 4·6 | 6,971 | 29 | 1·7 | 1·0, 2·9 | 10,071 | 18 | 1·2 | 0·6, 2·2 | 3,146 | 0 | 0·8 | 0·4, 1·8 |
| | <-3 | 1,722 | 59 | 8·2 | 8·2, 24·4 | 2,306 | 29 | 6·8 | 4·0, 11·4 | 3,011 | 13 | 3·3 | 1·7, 6·2 | 847 | 3 | 1·6 | 0·7, 3·7 |

[1] Risk = predicted absolute risk of dying within 1 month per 1,000 child months modelled in mixed linear regression models accounting for repeated measures in each child, clustering within studies, and time at risk

Abbreviations: WAZ = weight-for-age Z-score· Stunted was defined as height-for-age Z-score <-2·

**Table S6 - Absolute risk of death per 1,000 child months by age group and by individual exposures in Anthropometry-Selected (A-S) cohorts, i·e·, enrolment on the basis of anthropometric deficit**

| | <6 months | | | | 6-11 months | | | | 12-23 months | | | | 24-59 months | | | |
|---|---|---|---|---|---|---|---|---|---|---|---|---|---|---|---|---|
| **Indicator** | N | **Deaths** | **Risk[1]** | **95% CI** | N | **Deaths** | **Risk[1]** | **95% CI** | N | **Deaths** | **Risk[1]** | **95% CI** | N | **Deaths** | **Risk[1]** | **95% CI** |
| **ALL** | 10,767 | 39 | 3·0 | 0·0, 165·2 | 16,734 | 117 | 8·3 | 1·1, 60·4 | 23,090 | 140 | 5·4 | 0·7, 40·2 | 13,929 | 70 | 5·3 | 0·7, 39·5 |
| **WAZ** | | | | | | | | | | | | | | | | |
| ≥-2 | 4,390 | 1 | 0·8 | 0·1, 5·8 | 3,230 | 9 | 1·3 | 0·2, 7·8 | 3,740 | 8 | 2·0 | 0·3, 11·5 | 2,221 | 8 | 3·1 | 0·5, 19·3 |
| <-2, ≥-3 | 4,209 | 9 | 3·1 | 0·5, 18·5 | 4,561 | 19 | 3·5 | 0·6, 19·5 | 7,096 | 26 | 3·9 | 0·7, 21·5 | 4,050 | 15 | 4·3 | 0·7, 24·9 |
| <-3 | 1,414 | 28 | 19·4 | 3·5, 101 | 5,009 | 76 | 13·2 | 2·4, 69·2 | 9,242 | 100 | 8·9 | 1·6, 47·7 | 7,342 | 47 | 6·0 | 1·1, 33·3 |
| Missing | 754 | 1 | 2·1 | 0·1, 28·3 | 3,934 | 13 | 0·0 | 0, 1000 | 3,012 | 6 | 0·0 | 0, 1000 | 316 | 0 | 0·0 | 0, 1000 |
| **HAZ** | | | | | | | | | | | | | | | | |
| ≥-2 | 5,160 | 4 | 2·5 | 0·4, 15·8 | 5,281 | 21 | 2·7 | 0·5, 15·5 | 4,567 | 13 | 2·9 | 0·5, 16·4 | 3,094 | 7 | 3·0 | 0·5, 18·9 |
| <-2, ≥-3 | 3,277 | 20 | 10·9 | 1·9, 61·3 | 3,915 | 30 | 7·1 | 1·3, 39·1 | 5,936 | 26 | 4·6 | 0·8, 25·9 | 3,398 | 15 | 3·0 | 0·5, 17·8 |
| <-3 | 1,484 | 14 | 16·8 | 2·9, 90·0 | 3,589 | 53 | 11·4 | 2·0, 61·0 | 9,567 | 95 | 7·7 | 1·4, 41·7 | 7,117 | 48 | 5·2 | 0·9, 28·9 |
| Missing | 846 | 1 | 2·7 | 0·2, 37·4 | 3,949 | 13 | 0·0 | 0, 1000 | 3·020 | 6 | 0·0 | 0, 1000 | 320 | 0 | 0·0 | 0, 1000 |
| **WHZ** | | | | | | | | | | | | | | | | |
| ≥-2 | 7,889 | 12 | 2·7 | 0·4, 16·7 | 6,947 | 26 | 2·9 | 0·5, 17·3 | 10,427 | 31 | 3·2 | 0·5, 18·6 | 6,540 | 28 | 3·5 | 0·6, 20·7 |
| <-2, ≥-3 | 1,121 | 2 | 7·5 | 1·2, 45·5 | 4,034 | 33 | 6·1 | 1·0, 35·2 | 6,852 | 42 | 4·9 | 0·8, 28·4 | 4,781 | 15 | 4·0 | 0·6, 23·9 |
| <-3 | 330 | 13 | 25·7 | 4·3, 139 | 1,801 | 45 | 20·8 | 3·6, 112 | 2,789 | 61 | 16·7 | 2·9, 91·4 | 2,282 | 27 | 13·5 | 2·3, 76·5 |
| Missing | 1,427 | 12 | 13·4 | 2·0, 83·8 | 3,952 | 13 | 0·0 | 0, 1000 | 3,022 | 6 | 0·0 | 0, 1000 | 326 | 0 | 0·0 | 0, 1000 |
| **MUAC** | | | | | | | | | | | | | | | | |
| ≥125 | 3,272 | 1 | 1·5 | 0·2, 11·7 | 3,863 | 8 | 1·8 | 0·3, 12·5 | 8,082 | 20 | 2·1 | 0·3, 14·3 | 6,422 | 22 | 2·6 | 0·4, 17·5 |
| <125, ≥115 | 2,261 | 0 | 2·4 | 0·3, 17·2 | 8,511 | 38 | 3·0 | 0·4, 19·9 | 10,618 | 49 | 3·7 | 0·5, 24·0 | 5,620 | 18 | 4·5 | 0·7, 30·3 |
| <115 | 4,442 | 36 | 14·7 | 2·2, 92·4 | 4,113 | 71 | 13·0 | 2·0, 82·3 | 4,358 | 71 | 12·0 | 1·8, 74·9 | 1,858 | 30 | 10·8 | 1·6, 69·7 |
| Missing | 792 | 2 | 6·5 | 0·6, 66·7 | 247 | 0 | 0·0 | 0·0, 1000 | 32 | 0 | 0·0 | 0, 1000 | 29 | 0 | 0·0 | 0, 1000 |
| **MUAC (0-6 mo)** | | | | | | | | | | | | | | | | |
| ≥120 | 4,711 | 1 | 0·3 | 0·0, 1·7 | ·· | ·· | ·· | ·· | ·· | ·· | ·· | ·· | ·· | ·· | ·· | ·· |
| <120, ≥110 | 1,855 | 0 | 0·0 | 0·0, 1000 | ·· | ·· | ·· | ·· | ·· | ·· | ·· | ·· | ·· | ·· | ·· | ·· |
| <110 | 3,409 | 36 | 7·6 | 5·4, 10·6 | ·· | ·· | ·· | ·· | ·· | ·· | ·· | ·· | ·· | ·· | ·· | ·· |
| Missing | 792 | 2 | 2·6 | 0·6, 10·3 | ·· | ·· | ·· | ·· | ·· | ·· | ·· | ·· | ·· | ·· | ·· | ·· |
| **any BF** | | | | | | | | | | | | | | | | |
| No | 93 | 1 | 17·1 | 2·6, 105 | 533 | 15 | 10·6 | 1·7, 63·4 | 4,114 | 52 | 6·6 | 1·1, 39·0 | 6,356 | 56 | 4·1 | 0·7, 24·7 |
| Yes | 7,854 | 16 | 7·9 | 1·3, 47·2 | 9,440 | 87 | 5·9 | 1·0, 34·9 | 11,227 | 77 | 4·5 | 0·7, 26·7 | 1,319 | 11 | 3·4 | 0·5, 21·2 |
| Missing | 2,820 | 22 | 23·9 | 3·6, 1412 | 6,761 | 15 | 5·8 | 0·5, 64·8 | 7,749 | 11 | 1·4 | 0·0, 76·0 | 6,254 | 3 | 0·3 | 0·0, 111 |
| **LBW** | | | | | | | | | | | | | | | | |
| No | 1 | 0 | ·· | ·· | 2,135 | 20 | 9·3 | 6·2, 14·1 | 2,649 | 21 | 7·7 | 5·4, 10·9 | 592 | 3 | 6·3 | 3·1, 12·9 |
| Yes | 0 | 0 | ·· | ·· | 100 | 1 | 6·9 | 1·0, 48·6 | 134 | 1 | 11·0 | 3·5, 33·9 | 41 | 1 | 17·4 | 2·4, 116·3 |
| Missing | 0 | 0 | ·· | ·· | 276 | 3 | 9·6 | 3·4, 26·8 | 390 | 4 | 10·6 | 5·4, 20·5 | 161 | 2 | 11·7 | 3·5, 38·3 |
| **PTB** | | | | | | | | | | | | | | | | |
| No | 0 | ·· | ·· | ·· | 0 | ·· | ·· | ·· | 0 | ·· | ·· | ·· | 0 | ·· | ·· | ·· |
| Yes | 0 | ·· | ·· | ·· | 0 | ·· | ·· | ·· | 0 | ·· | ·· | ·· | 0 | ·· | ·· | ·· |
| Missing | 0 | ·· | ·· | ·· | 0 | ·· | ·· | ·· | 0 | ·· | ·· | ·· | 0 | ·· | ·· | ·· |
| **Diarrhea** | | | | | | | | | | | | | | | | |
| No | 8,119 | 16 | 4·1 | 0·6, 26·4 | 8,270 | 32 | 3·6 | 0·6, 22·3 | 12,153 | 48 | 3·1 | 0·5, 19·5 | 8,629 | 31 | 2·7 | 0·4,17·7 |
| Yes | 425 | 2 | 13·5 | 2·1, 82·0 | 4,531 | 74 | 10·6 | 1·7, 63·8 | 7,176 | 86 | 8·3 | 1·3, 50·5 | 3,976 | 38 | 6·5 | 1·0, 40·6 |
| Missing | 0 | 0 | ·· | ·· | 3,910 | 11 | 0·0 | 0, 1000 | 3,761 | 6 | 0·0 | 0, 1000 | 1,324 | 1 | 55·3 | 3·5, 494 |

| | | | | | | | | | | | | | | | | |
|---|---|---|---|---|---|---|---|---|---|---|---|---|---|---|---|---|
| **LRTI** | | | | | | | | | | | | | | | | |
| No | 0 | ·· | ·· | ·· | 0 | ·· | ·· | ·· | 0 | ·· | ·· | ·· | 0 | ·· | ·· | ·· |
| Yes | 0 | ·· | ·· | ·· | 0 | ·· | ·· | ·· | 0 | ·· | ·· | ·· | 0 | ·· | ·· | ·· |
| Missing | 0 | ·· | ·· | ·· | 0 | ·· | ·· | ·· | 0 | ·· | ·· | ·· | 0 | ·· | ·· | ·· |
| **Severe illness**[2] | | | | | | | | | | | | | | | | |
| No | 5 | 0 | ·· | ·· | 7,957 | 66 | 6·4 | 0·3, 122 | 14,403 | 94 | 4·5 | 0·2, 89 | 10,628 | 46 | 3·2 | 0·1, 65·3 |
| Yes | 0 | 0 | ·· | ·· | 935 | 12 | 11·6 | 0·5, 210 | 1,511 | 14 | 12·5 | 0·6, 216 | 1,037 | 17 | 13·5 | 0·6, 233 |

[1] Risk = predicted absolute risk of dying within 1 month per 1,000 child months modelled in mixed linear regression models accounting for repeated measures in each child, clustering within studies, and time at risk

[2] Severe illness was defined as having any of the following: being admitted to hospital, not being able to drink or breastfeed, having had convulsions, being lethargic or unconscious·

Abbreviations: BF = breastfeeding; HAZ = height-for-age Z-score; LBW= low birthweight; MUAC = mid-upper arm circumference; PTB = preterm birth; WAZ = weight-for-age Z-score; WHZ= weight-for-height Z-score

**Table S7 - Mortality odds ratio by age group and by individual exposures in Anthropometry-Selected (A-S) cohorts, i·e·, enrolment on the basis of anthropometric deficit**

| | <6 months | | | | 6-11 months | | | | 12-23 months | | | | 24-59 months | | | |
|---|---|---|---|---|---|---|---|---|---|---|---|---|---|---|---|---|
| **Indicator** | **N** | **Deaths** | **OR** | **95% CI** | **N** | **Deaths** | **OR** | **95% CI** | **N** | **Deaths** | **OR** | **95% CI** | **N** | **Deaths** | **OR** | **95% CI** |
| **WAZ** | | | | | | | | | | | | | | | | |
| ≥-2 | 4,390 | 1 | Ref | | 3,230 | 9 | Ref | | 3,740 | 8 | Ref | | 2,221 | 8 | Ref | |
| <-2, ≥-3 | 4,209 | 9 | 0·93 | 0·11, 7·67 | 4,561 | 19 | 3·30 | 0·01, 767 | 7,096 | 26 | 1·96 | 0·02, 203 | 4,050 | 15 | 1·83 | 0·05, 63·1 |
| <-3 | 1,414 | 28 | 7·99 | 1·05, 60·8 | 5,009 | 76 | 9·55 | 0·07, 1271 | 9,242 | 100 | 13·4 | 0·18, 972 | 7,342 | 47 | 30·3 | 1·27, 722 |
| Missing | 754 | 1 | 0·42 | 0·0, 150 | 3,934 | 13 | ·· | ·· | 3,012 | 6 | ·· | ·· | 316 | 0 | ·· | ·· |
| **HAZ** | | | | | | | | | | | | | | | | |
| ≥-2 | 5,160 | 4 | Ref | | 5,281 | 21 | Ref | | 4,567 | 13 | Ref | | 3,094 | 7 | Ref | |
| <-2, ≥-3 | 3,277 | 20 | 8·10 | 2·11, 31·1 | 3,915 | 30 | 0·18 | 0·00, 6·64 | 5,936 | 26 | 0·28 | 0·01, 15·2 | 3,398 | 15 | 1·40 | 0·04, 55·2 |
| <-3 | 1,484 | 14 | 10·6 | 1·39, 42·4 | 3,589 | 53 | 6·03 | 0·23, 159 | 9,567 | 95 | 2·16 | 0·06, 79·4 | 7,117 | 48 | 4·74 | 0·18, 122 |
| Missing | 846 | 1 | 0·74 | 0·0, 198 | 3,949 | 13 | ·· | ·· | 3·020 | 6 | ·· | ·· | 320 | 0 | ·· | ·· |
| **WHZ** | | | | | | | | | | | | | | | | |
| ≥-2 | 7,889 | 12 | Ref | | 6,947 | 26 | Ref | | 10,427 | 31 | Ref | | 6,540 | 28 | Ref | |
| <-2, ≥-3 | 1,121 | 2 | 0·59 | 0·12, 2·89 | 4,034 | 33 | 4·53 | 0·20, 103 | 6,852 | 42 | 14·1 | 1·12, 178 | 4,781 | 15 | 0·46 | 0·03, 6·43 |
| <-3 | 330 | 13 | 4·72 | 1·53, 14·6 | 1,801 | 45 | 2·93 | 0·15, 58·2 | 2,789 | 61 | 6·91 | 0·69, 69·3 | 2,282 | 27 | 33·0 | 2·68, 407 |
| Missing | 1,427 | 12 | 2·65 | 1·00, 6·99 | 3,952 | 13 | ·· | ·· | 3,022 | 6 | ·· | ·· | 326 | 0 | ·· | ·· |
| **MUAC** | | | | | | | | | | | | | | | | |
| ≥125 | 3,272 | 1 | Ref | | 3,863 | 8 | Ref | | 8,082 | 20 | Ref | | 6,422 | 22 | Ref | |
| <125, ≥115 | 2,261 | 0 | ·· | ·· | 8,511 | 38 | 1·71 | 0·01, 273 | 10,618 | 49 | 4·06 | 0·23, 72·0 | 5,620 | 18 | 16·6 | 0·86, 326 |
| <115 | 4,442 | 36 | 1·66 | 0·0, 1334 | 4,113 | 71 | 5·13 | 0·05, 590 | 4,358 | 71 | 4·01 | 0·27, 58·6 | 1,858 | 30 | 13·5 | 1·17, 156 |
| Missing | 792 | 2 | 2·20 | 0·0, 3427 | 247 | 0 | ·· | ·· | 32 | 0 | ·· | ·· | 29 | 0 | ·· | ·· |
| **MUAC (0-6 mo)** | | | | | | | | | | | | | | | | |
| ≥120 | 4,711 | 1 | Ref | | | | | | | | | | | | | |
| <120, ≥110 | 1,855 | 0 | ·· | ·· | ·· | ·· | ·· | ·· | ·· | ·· | ·· | ·· | ·· | ·· | ·· | ·· |
| <110 | 3,409 | 36 | 7·87 | 0·02,3703 | ·· | ·· | ·· | ·· | ·· | ·· | ·· | ·· | ·· | ·· | ·· | ·· |
| Missing | 792 | 2 | 10·2 | 0·01,9830 | ·· | ·· | ·· | ·· | ·· | ·· | ·· | ·· | ·· | ·· | ·· | ·· |
| **any BF** | | | | | | | | | | | | | | | | |
| No | 93 | 1 | Ref | | 533 | 15 | Ref | | 4,114 | 52 | Ref | | 6,356 | 56 | Ref | |
| Yes | 7,854 | 16 | ·· | ·· | 9,440 | 87 | 0·85 | 0·03, 23·2 | 11,227 | 77 | 0·24 | 0·03, 1·74 | 1,319 | 11 | 1·11 | 0·06, 21·6 |
| Missing | 2,820 | 22 | ·· | ·· | 6,761 | 15 | ·· | ·· | 7,749 | 11 | ·· | ·· | 6,254 | 3 | ·· | ·· |
| **LBW** | | | | | | | | | | | | | | | | |
| No | 1 | 0 | Ref | | 2,135 | 20 | Ref | | 2,649 | 21 | Ref | | 592 | 3 | Ref | |
| Yes | 0 | 0 | ·· | ·· | 100 | 1 | ·· | ·· | 134 | 1 | 0·22 | 0·0, 3975 | 41 | 1 | ·· | ·· |
| Missing | 0 | 0 | ·· | ·· | 276 | 3 | 0·46 | 0·0, 646 | 390 | 4 | ·· | ·· | 161 | 2 | ·· | ·· |
| **PTB** | | | | | | | | | | | | | | | | |
| No | 0 | ·· | ·· | ·· | 0 | ·· | ·· | ·· | 0 | ·· | ·· | ·· | 0 | ·· | ·· | ·· |
| Yes | 0 | ·· | ·· | ·· | 0 | ·· | ·· | ·· | 0 | ·· | ·· | ·· | 0 | ·· | ·· | ·· |
| Missing | 0 | ·· | ·· | ·· | 0 | ·· | ·· | ·· | 0 | ·· | ·· | ·· | 0 | ·· | ·· | ·· |
| **Diarrhea** | | | | | | | | | | | | | | | | |
| No | 8,119 | 16 | Ref | | 8,270 | 32 | Ref | | 12,153 | 48 | Ref | | 8,629 | 31 | Ref | |
| Yes | 425 | 2 | 0·66 | ·· | 4,531 | 74 | 12·7 | 0·96, 169 | 7,176 | 86 | 4·70 | 0·70, 31·8 | 3,976 | 38 | 0·80 | 0·10, 6·69 |
| Missing | 0 | 0 | ·· | ·· | 3,910 | 11 | ·· | ·· | 3,761 | 6 | ·· | ·· | 1,324 | 1 | ·· | ·· |

| | | | | | | | | | | | | | | | | |
|---|---|---|---|---|---|---|---|---|---|---|---|---|---|---|---|---|
| **LRTI** | | | | | | | | | | | | | | | | |
| No | 0 | ·· | ·· | ·· | 0 | ·· | ·· | ·· | 0 | ·· | ·· | ·· | 0 | ·· | ·· | ·· |
| Yes | 0 | ·· | ·· | ·· | 0 | ·· | ·· | ·· | 0 | ·· | ·· | ·· | 0 | ·· | ·· | ·· |
| Missing | 0 | ·· | ·· | ·· | 0 | ·· | ·· | ·· | 0 | ·· | ·· | ·· | 0 | ·· | ·· | ·· |
| **Severe illness[1]** | | | | | | | | | | | | | | | | |
| No | 5 | 0 | ·· | ·· | 7,957 | 66 | Ref | | 14,403 | 94 | Ref | | 10,628 | 46 | Ref | |
| Yes | 0 | 0 | ·· | ·· | 935 | 12 | 13·2 | 0·30, 591 | 1,511 | 14 | 0·55 | 0·02, 12·4 | 1,037 | 17 | 3·81 | 0·34, 42·8 |

[1] Severe illness was defined as having any of the following: being admitted to hospital, not being able to drink or breastfeed, having had convulsions, being lethargic or unconscious·

Abbreviations: BF = breastfeeding; HAZ = height-for-age Z-score; LBW= low birthweight; MUAC = mid-upper arm circumference; PTB = preterm birth; WAZ = weight-for-age Z-score; WHZ= weight-for-height Z-score

**Table S8– Absolute risk of death per 1,000 child months by age group and by individual exposures in Illness-Selected (I-S) cohorts, i·e·, enrolment on the basis of the presence of illness**

| | <6 months | | | | 6-11 months | | | | 12-23 months | | | | 24-59 months | | | |
|---|---|---|---|---|---|---|---|---|---|---|---|---|---|---|---|---|
| **Indicator** | N | **Deaths** | **Risk[1]** | **95% CI** | N | | **Risk[1]** | **95% CI** | N | | **Risk[1]** | **95% CI** | N | | **Risk[1]** | **95% CI** |
| **ALL** | 3,625 | 458 | 12·3 | 9·2, 16·3 | 4,999 | 285 | 7·8 | 5·7, 10·6 | 7,676 | 312 | 6·9 | 5·1, 9·2 | 2,434 | 146 | 5·3 | 3·5, 8·1 |
| **WAZ** | | | | | | | | | | | | | | | | |
| ≥-2 | 2,324 | 188 | 3·1 | 0·9, 10·3 | 2,639 | 82 | 3·4 | 1·1, 10·6 | 3,886 | 94 | 3·7 | 1·2, 11·8 | 1,563 | 85 | 4·1 | 1·2, 14·2 |
| <-2, ≥-3 | 493 | 86 | 14·3 | 3·8, 51·4 | 878 | 37 | 10·5 | 3·2, 33·9 | 1,604 | 53 | 7·7 | 2·3, 25·6 | 449 | 25 | 5·6 | 1·4, 22·1 |
| <-3 | 780 | 182 | 46·0 | 13·7,143 | 1,246 | 157 | 40·3 | 12·9,119 | 1,752 | 152 | 35·2 | 11·0,107 | 341 | 32 | 30·8 | 8·5, 105 |
| Missing | 28 | 2 | 71·4 | 9·6, 378 | 236 | 9 | 58·5 | 14·1, 212 | 434 | 13 | 47·8 | 11·8, 174 | 81 | 4 | 38·9 | 5·5, 229 |
| **HAZ** | | | | | | | | | | | | | | | | |
| ≥-2 | 2,486 | 243 | 5·1 | 2·2, 11·8 | 2,908 | 126 | 4·4 | 2·1, 9·4 | 3,491 | 116 | 3·8 | 1·8, 8·3 | 1,486 | 77 | 3·3 | 1·3, 8·2 |
| <-2, ≥-3 | 490 | 85 | 20·6 | 7·7, 53·8 | 902 | 59 | 14·2 | 6·3, 31·9 | 1,860 | 66 | 9·8 | 4·2, 22·7 | 457 | 26 | 6·7 | 2·3, 19·2 |
| <-3 | 610 | 126 | 31·0 | 12·2, 76·6 | 940 | 89 | 25·7 | 11·6, 56·2 | 1,875 | 109 | 21·3 | 9·3, 47·9 | 403 | 37 | 17·7 | 6·5, 47·2 |
| Missing | 39 | 4 | 38·6 | 6·7, 192 | 249 | 11 | 42·0 | 13·7, 122 | 450 | 21 | 45·6 | 15·7, 126 | 88 | 6 | 49·6 | 9·7, 218 |
| **WHZ** | | | | | | | | | | | | | | | | |
| ≥-2 | 2,435 | 242 | 6·5 | 3·4, 12·4 | 3,194 | 103 | 5·4 | 3·1, 9·4 | 5,003 | 113 | 4·5 | 2·5, 8·0 | 1,754 | 89 | 3·8 | 1·8, 7·6 |
| <-2, ≥-3 | 461 | 64 | 12·6 | 5·4, 28·8 | 802 | 60 | 10·6 | 5·6, 20·1 | 1,230 | 53 | 8·9 | 4·5, 17·6 | 326 | 21 | 7·5 | 3·0, 18·8 |
| <-3 | 644 | 132 | 21·2 | 10·2, 43·6 | 753 | 111 | 23·2 | 12·8, 41·8 | 992 | 125 | 25·4 | 13·3, 48·0 | 258 | 26 | 27·7 | 11·8, 63·7 |
| Missing | 85 | 29 | 40·7 | 11·1, 139 | 250 | 11 | 41·6 | 17·7, 94·6 | 451 | 21 | 42·5 | 17·4, 100 | 96 | 10 | 43·4 | 11·0, 157 |
| **MUAC** | | | | | | | | | | | | | | | | |
| ≥125 | 1,019 | 52 | 3·0 | 1·3, 7·1 | 2,863 | 81 | 3·2 | 1·5, 6·7 | 5,184 | 121 | 3·3 | 1·6, 6·9 | 2,161 | 118 | 3·5 | 1·5, 7·9 |
| <125, ≥115 | 735 | 70 | 9·4 | 3·7, 23·6 | 997 | 56 | 11·4 | 5·2, 24·7 | 1,264 | 57 | 13·9 | 5·8, 32·9 | 123 | 10 | 17·0 | 5·4, 52·4 |
| <115 | 1,871 | 333 | 22·9 | 10·2, 50·7 | 905 | 141 | 41·8 | 20·2, 84·5 | 798 | 122 | 75·0 | 33·1,161 | 68 | 17 | 131 | 47·5, 313 |
| Missing | 31 | 3 | 63·5 | 10·8, 297 | 234 | 7 | 47·2 | 15·4, 135 | 430 | 12 | 34·9 | 11·6, 101 | 82 | 1 | 25·8 | 4·4, 137 |
| **MUAC (0-6 mo)** | | | | | | | | | | | | | | | | |
| ≥120 | 1,371 | 92 | 4·7 | 2·6, 8·5 | ·· | ·· | ·· | ·· | ·· | ·· | ·· | ·· | ·· | ·· | ·· | ·· |
| <120, ≥110 | 829 | 76 | 6·4 | 3·2, 12·9 | ·· | ·· | ·· | ·· | ·· | ·· | ·· | ·· | ·· | ·· | ·· | ·· |
| <110 | 1,394 | 287 | 31·9 | 20·4, 49·4 | ·· | ·· | ·· | ·· | ·· | ·· | ·· | ·· | ·· | ·· | ·· | ·· |
| Missing | 31 | 3 | 90·3 | 6·3, 608 | ·· | ·· | ·· | ·· | ·· | ·· | ·· | ·· | ·· | ·· | ·· | ·· |
| **any BF** | | | | | | | | | | | | | | | | |
| No | 319 | 44 | 26·0 | 11·6, 57·2 | 996 | 96 | 16·9 | 9·2, 31·0 | 2,726 | 179 | 11·0 | 6·5, 18·6 | 2,015 | 144 | 7·2 | 3·9, 13·2 |
| Yes | 3,303 | 413 | 9·0 | 5·0, 16·1 | 4,002 | 189 | 6·7 | 4·1, 11·1 | 4,949 | 132 | 5·1 | 2·8, 9·3 | 417 | 2 | 3·8 | 1·6, 9·0 |
| Missing | 3 | 1 | ·· | ·· | 1 | 0 | ·· | ·· | 1 | 1 | ·· | ·· | 2 | 0 | ·· | ·· |
| **LBW** | | | | | | | | | | | | | | | | |
| No | 3,014 | 372 | 10·5 | 7·9, 13·9 | 2,966 | 163 | 7·9 | 6·5, 9·7 | 4,686 | 190 | 6·0 | 4·7, 7·7 | 1,610 | 103 | 4·5 | 3·1, 6·7 |
| Yes | 291 | 51 | 17·8 | 7·1, 43·6 | 565 | 27 | 14·4 | 7·8, 26·5 | 867 | 23 | 11·6 | 5·2, 26·0 | 116 | 5 | 9·4 | 2·6, 33·9 |
| Missing | 296 | 31 | 20·2 | 7·6, 52·8 | 1,099 | 50 | 14·1 | 8·0, 24·7 | 1,722 | 55 | 9·8 | 5·1, 18·9 | 229 | 1 | 6·9 | 2·2, 21·4 |
| **PTB** | | | | | | | | | | | | | | | | |
| No | 2,473 | 344 | 7·9 | 1·9, 31·1 | 1,082 | 90 | 7·6 | 2·0, 28·4 | 1,366 | 127 | 7·3 | 1·8, 28·8 | 1,172 | 101 | 7·0 | 1·5, 32·2 |
| Yes | 166 | 39 | 23·0 | 3·8, 127 | 48 | 5 | 15·5 | 2·8, 81·8 | 33 | 5 | 10·4 | 1·0, 96·7 | 27 | 2 | 7·0 | 0·3, 151·2 |
| Missing | 5 | 1 | ·· | ·· | 10 | 4 | ·· | ·· | 15 | 1 | ·· | ·· | 17 | 2 | ·· | ·· |
| **Diarrhea** | | | | | | | | | | | | | | | | |
| No | 2,637 | 344 | 7·7 | 2·4, 24·5 | 2,279 | 138 | 6·8 | 2·2, 20·6 | 3,817 | 163 | 6·0 | 2·0, 18·4 | 1,780 | 96 | 5·3 | 1·6, 17·5 |
| Yes | 986 | 112 | 8·3 | 2·5, 27·5 | 2,705 | 132 | 6·9 | 2·2, 21·5 | 3,836 | 126 | 5·8 | 1·8, 18·5 | 632 | 28 | 4·8 | 1·3, 17·4 |
| Missing | 0 | 0 | ·· | ·· | 0 | 0 | ·· | ·· | 0 | 0 | ·· | ·· | 0 | 0 | ·· | ·· |
| **LRTI** | | | | | | | | | | | | | | | | |

| | | | | | | | | | | | | | | | | |
|---|---|---|---|---|---|---|---|---|---|---|---|---|---|---|---|---|
| No | 2,177 | 264 | 6·6 | 2·0, 21·0 | 2,609 | 136 | 5·7 | 1·9, 17·7 | 4,621 | 172 | 4·9 | 1·6, 15·1 | 1,782 | 76 | 4·3 | 1·3, 14·1 |
| Yes | 1,445 | 192 | 9·5 | 2·9, 30·5 | 2,375 | 134 | 9·0 | 2·9, 27·1 | 3,032 | 117 | 8·5 | 2·7, 26·2 | 630 | 48 | 8·0 | 2·3, 27·5 |
| Missing | 3 | 2 | - | - | 15 | 15 | - | - | 23 | 23 | 1000 | 0, 1000 | 22 | 22 | 1000 | 0, 1000 |
| **Severe illness[2]** | | | | | | | | | | | | | | | | |
| No | 2,814 | 279 | 5·7 | 2·5, 13·1 | 3,295 | 135 | 5·5 | 2·6, 12·0 | 5,019 | 143 | 5·4 | 2·4, 12·1 | 1,383 | 70 | 5·3 | 2·1, 13·5 |
| Yes | 787 | 175 | 34·2 | 14·0, 81·3 | 1,335 | 105 | 25·4 | 11·4, 55·6 | 2,256 | 125 | 18·8 | 8·1, 42·9 | 572 | 39 | 13·9 | 5·1, 37·1 |
| **Nr· of severe illnesses[2]** | | | | | | | | | | | | | | | | |
| 0 | 2,814 | 279 | 5·8 | 2·6, 13·1 | 3,295 | 135 | 5·6 | 2·7, 11·9 | 5,019 | 143 | 5·5 | 2·5, 12·0 | 1,383 | 70 | 5·3 | 2·1, 13·3 |
| 1 | 654 | 136 | 28·4 | 11·4, 69·0 | 1,121 | 76 | 21·0 | 9·4, 46·2 | 1,796 | 92 | 15·6 | 6·7, 35·9 | 456 | 26 | 11·5 | 4·1, 31·8 |
| ≥ 2 | 133 | 39 | 83·9 | 22·8, 265 | 214 | 29 | 57·3 | 20·1, 146 | 460 | 33 | 38·7 | 13·4, 107 | 116 | 13 | 26·0 | 6·0, 106 |

[1] Risk = predicted absolute risk of dying within 1 month per 1,000 child months modelled in mixed linear regression models accounting for repeated measures in each child, clustering within studies, and time at risk

[2] Severe illness was defined as having any of the following: being admitted to hospital, not being able to drink or breastfeed, having had convulsions, being lethargic or unconscious·

Abbreviations: BF = breastfeeding; HAZ = height-for-age Z-score; LBW= low birthweight; MUAC = mid-upper arm circumference; PTB = preterm birth; WAZ = weight-for-age Z-score; WHZ= weight-for-height Z-score

**Table S9 - Mortality odds ratio by age group and by individual exposures in Illness-Selected(I-S) cohorts, i·e·, enrolment on the basis of the presence of illness**

| | <6 months | | | | 6-11 months | | | | 12-23 months | | | | 24-59 months | | | |
|---|---|---|---|---|---|---|---|---|---|---|---|---|---|---|---|---|
| **Indicator** | N | **Deaths** | **OR** | **95% CI** | N | **Deaths** | **OR** | **95% CI** | N | **Deaths** | **OR** | **95% CI** | N | **Deaths** | **OR** | **95% CI** |
| **WAZ** | | | | | | | | | | | | | | | | |
| ≥-2 | 2,324 | 188 | Ref | | 2,639 | 82 | Ref | | 3,886 | 94 | Ref | | 1,563 | 85 | Ref | |
| <-2, ≥-3 | 493 | 86 | 8·38 | 1·81, 38·5 | 878 | 37 | 14·4 | 0·07, | 1,604 | 53 | 1·83 | 0·02, 180 | 449 | 25 | 2·27 | 0·02, 253 |
| <-3 | 780 | 182 | 15·7 | 3·95, 62·3 | 1,246 | 157 | 5·00· | 2898 | 1,752 | 152 | 1·30 | 0·03, 55·4 | 341 | 32 | 14·8 | 0·13, |
| Missing | 28 | 2 | ·· | ·· | 236 | 9 | 0·18 | 0·13, 205 | 434 | 13 | 0·30 | 0·0, 331 | 81 | 4 | ·· | 1705 |
| | | | | | | | | 0·0, 895 | | | | | | | | ·· |
| **HAZ** | | | | | | | | | | | | | | | | |
| ≥-2 | 2,486 | 243 | Ref | | 2,908 | 126 | Ref | | 3,491 | 116 | Ref | | 1,486 | 77 | Ref | |
| <-2, ≥-3 | 490 | 85 | 5·09 | 1·05, 24·7 | 902 | 59 | 5·64 | 0·07, 450 | 1,860 | 66 | 2·05 | 0·04, 118 | 457 | 26 | 0·90 | 0·01, 111 |
| <-3 | 610 | 126 | 7·63 | 1·44, 40·4 | 940 | 89 | 1·36 | 0·03, 71·0 | 1,875 | 109 | 1·01 | 0·02, 48·5 | 403 | 37 | 2·55 | 0·03, 228 |
| Missing | 39 | 4 | ·· | ·· | 249 | 11 | 0·33 | 0·0, 788 | 450 | 21 | 0·44 | 0·0, 245 | 88 | 6 | 0·0 | 0·0, 145 |
| **WHZ** | | | | | | | | | | | | | | | | |
| ≥-2 | 2,435 | 242 | Ref | | 3,194 | 103 | Ref | | 5,003 | 113 | Ref | | 1,754 | 89 | Ref | |
| <-2, ≥-3 | 461 | 64 | 1·52 | 0·31, 7·40 | 802 | 60 | 96·2 | 0·97, | 1,230 | 53 | 0·07 | 0·00, 4·74 | 326 | 21 | 1·38 | 0·01, 139 |
| <-3 | 644 | 132 | 1·94 | 0·58, 6·48 | 753 | 111 | 2·24 | 9584 | 992 | 125 | 4·19 | 0·11, 167 | 258 | 26 | 41·8 | 0·28, |
| Missing | 85 | 29 | 13·8 | 1·05, 180 | 250 | 11 | 0·51 | 0·04, 119 | 451 | 21 | 0·45 | 0·0, 203 | 96 | 10 | 0·03 | 6268 |
| | | | | | | | | 0·0, 1063 | | | | | | | | 0·0, 92·1 |
| **MUAC** | | | | | | | | | | | | | | | | |
| ≥125 | 1,019 | 52 | Ref | | 2,863 | 81 | Ref | | 5,184 | 121 | Ref | | 2,161 | 118 | Ref | |
| <125, ≥115 | 735 | 70 | 2·63 | 0·19, 36·1 | 997 | 56 | 13·0 | 0·15, | 1,264 | 57 | 0·07 | 0·00, 4·28 | 123 | 10 | ·· | ·· |
| <115 | 1,871 | 333 | 9·57 | 1·00, 91·9 | 905 | 141 | 10·2 | 1138 | 798 | 122 | 11·8 | 0·27, 514 | 68 | 17 | 0·14 | 0·00, |
| Missing | 31 | 3 | ·· | ·· | 234 | 7 | 0·12 | 0·23, 458 | 430 | 12 | 0·71 | 0·0, 505 | 82 | 1 | ·· | 6145 |
| | | | | | | | | 0·0, 1125 | | | | | | | | ·· |
| **MUAC (0-6 mo)** | | | | | | | | | | | | | | | | |
| ≥120 | 1,371 | 92 | Ref | | ·· | ·· | ·· | ·· | ·· | ·· | ·· | ·· | ·· | ·· | ·· | ·· |
| <120, ≥110 | 829 | 76 | 1·02 | 0·15, 6·90 | ·· | ·· | ·· | ·· | ·· | ·· | ·· | ·· | ·· | ·· | ·· | ·· |
| <110 | 1,394 | 287 | 5·75 | 1·13, 29·2 | ·· | ·· | ·· | ·· | ·· | ·· | ·· | ·· | ·· | ·· | ·· | ·· |
| Missing | 31 | 3 | ·· | ·· | ·· | ·· | ·· | ·· | ·· | ·· | ·· | ·· | ·· | ·· | ·· | ·· |
| **any BF** | | | | | | | | | | | | | | | | |
| No | 319 | 44 | Ref | | 996 | 96 | Ref | | 2,726 | 179 | Ref | | 2,015 | 144 | Ref | |
| Yes | 3,303 | 413 | 0·94 | 0·06, 15·0 | 4,002 | 189 | 1·22 | 0·03, 43·8 | 4,949 | 132 | 0·12 | 0·01, 2·54 | 417 | 2 | ·· | ·· |
| Missing | 3 | 1 | ·· | ·· | 1 | 0 | ·· | ·· | 1 | 1 | ·· | ·· | 2 | 0 | ·· | ·· |
| **LBW** | | | | | | | | | | | | | | | | |
| No | 3,014 | 372 | Ref | | 2,966 | 163 | Ref | | 4,686 | 190 | Ref | | 1,610 | 103 | Ref | |
| Yes | 291 | 51 | 9·36 | 1·24, 70·6 | 565 | 27 | 3·67 | 0·02, 673 | 867 | 23 | 3·38 | 0·01, 848 | 116 | 5 | ·· | ·· |
| Missing | 296 | 31 | 3·93 | 0·07, 222 | 1,099 | 50 | 0·03 | 0·00, 2·19 | 1,722 | 55 | 0·76 | 0·01, 40·6 | 229 | 1 | ·· | ·· |
| **PTB** | | | | | | | | | | | | | | | | |
| No | 2,473 | 344 | Ref | | 1,082 | 90 | Ref | | 1,366 | 127 | Ref | | 1,172 | 101 | Ref | |
| Yes | 166 | 39 | 3·26 | 0·31, 33·7 | 48 | 5 | 0·01 | ·· | 33 | 5 | 0·18 | ·· | 27 | 2 | ·· | ·· |
| Missing | 5 | 1 | ·· | ·· | 10 | 4 | ·· | ·· | 15 | 1 | ·· | ·· | 17 | 2 | ·· | ·· |
| **Diarrhea** | | | | | | | | | | | | | | | | |

| | | | | | | | | | | | | | | | | |
|---|---|---|---|---|---|---|---|---|---|---|---|---|---|---|---|---|
| No | 2,637 | 344 | Ref | | 2,279 | 138 | Ref | | 3,817 | 163 | Ref | | 1,780 | 96 | Ref | |
| Yes | 986 | 112 | 1·95 | 0·41, 9·25 | 2,705 | 132 | 0·41 | 0·02, 9·16 | 3,836 | 126 | 1·07 | 0·05, 22·3 | 632 | 28 | 0·14 | 0·0, 10·4 |
| Missing | 0 | 0 | ·· | ·· | 0 | 0 | ·· | ·· | 0 | 0 | ·· | ·· | 0 | 0 | ·· | ·· |
| **LRTI** | | | | | | | | | | | | | | | | |
| No | 2,177 | 264 | Ref | | 2,609 | 136 | Ref | | 4,621 | 172 | Ref | | 1,782 | 76 | Ref | |
| Yes | 1,445 | 192 | 2·37 | 0·70, 7·97 | 2,375 | 134 | 0·99 | 0·04, 22·6 | 3,032 | 117 | 0·43 | 0·02, 9·97 | 630 | 48 | 0·33 | 0·01, 15·7 |
| Missing | 3 | 2 | ·· | ·· | 15 | 15 | ·· | ·· | 23 | 23 | ·· | ·· | 22 | 22 | ·· | ·· |
| **Severe illness**[1] | | | | | | | | | | | | | | | | |
| No | 2,814 | 279 | Ref | | 3,295 | 135 | Ref | | 5,019 | 143 | Ref | | 1,383 | 70 | Ref | |
| Yes | 787 | 175 | 22·2 | 7·26, 67·8 | 1,335 | 105 | 2·0 | 0·07, 60·3 | 2,256 | 125 | 1·45 | 0·05, 39·5 | 572 | 39 | 2·04 | 0·04, 112 |
| **Nr of severe illnesses**[1] | | | | | | | | | | | | | | | | |
| 0 | 2,814 | 279 | Ref | | 3,295 | 135 | Ref | | 5,019 | 143 | Ref | | 1,383 | 70 | Ref | |
| 1 | 654 | 136 | 19·0 | 5·64, 64·1 | 1,121 | 76 | 4·50 | 0·1, 194 | 1,796 | 92 | 0·97 | 0·03, 35·4 | 456 | 26 | 0·70 | 0·01, 68·2 |
| ≥ 2 | 133 | 39 | 39·1 | 4·75, 321 | 214 | 29 | 0·28 | 0·0, 98·8 | 460 | 33 | 7·38 | 0·02, 2355 | 116 | 13 | 53·7 | ·· |

[1] Severe illness was defined as having any of the following: being admitted to hospital, not being able to drink or breastfeed, having had convulsions, being lethargic or unconscious·

Abbreviations: BF = breastfeeding; HAZ = height-for-age Z-score; LBW= low birthweight; MUAC = mid-upper arm circumference; PTB = preterm birth; WAZ = weight-for-age Z-score; WHZ= weight-for-height Z-score
